# Design and development of a urinary cell pellet mRNA PCR-based assay for progressive kidney disease: Nephro-Dx

**DOI:** 10.1101/2025.10.31.25338655

**Authors:** Ashwani Kumar, Gabriel Barsotti, Zhengzi Yi, Zeguo Sun, Anand Reghuvaran, EM Tanvir, John Pell, Hongmei Shi, Sudhir Perincheri, Melissa Shaw, Candice Kent, Daris Javed, Petra Leite, Deepa Jayaram, Jeffrey Turner, Kristin Meliambro, Randy Luciano, John Cijiang He, Dennis Moledina, Perry Wilson, Weijia Zhang, Madhav C Menon

**Affiliations:** Section of Nephrology, Department of Medicine, Yale University School of Medicine, New Haven, Connecticut, USA; Division of Nephrology, Department of Medicine, Icahn School of Medicine at Mount Sinai, New York, New York, USA; Department of Pathology, Yale School of Medicine, New Haven, Connecticut, USA

## Abstract

Progressive chronic kidney disease (CKD) is a major source of public health spending. Current non-invasive tests estimate CKD but provide a minimal understanding of cell- or compartment-specific injury. The gold standard for CKD diagnosis is a kidney biopsy, which affords risks and is impractical to repeat multiple times. Hence, repeatable, non-invasive tests to estimate pathologic kidney injury for diagnoses, prognosis and follow-up of CKD represent a knowledge gap. We hypothesized that urinary shedding of specific cells is proportional to injury of those cells on biopsy, and that tracking cell-specific urine mRNA will correlate with ongoing injury. Informed by apriori biopsy and urine single cell RNA studies, we developed a targeted 10-gene urine mRNA assay to estimate kidney injury non-invasively (Nephro-Dx). In a pilot study of 48 patients with diverse kidney pathology on biopsy and 20 controls, we confirmed our assay’s utility in differentiating any kidney disease from controls. Within biopsied cases, we confirmed correlations of cell-specific urinary gene expression with corresponding compartment injury on biopsy using a validated quantitative digital pathology platform. We show that the gene signatures including individual genes associate with subsequent loss of kidney function within our cases providing an advantage over existing non-invasive tests. Our parsimonious set of gene signatures in Nephro-Dx shows advantages in early diagnosis, monitoring, and prognosis to impact this public health problem.

## Introduction

Kidney function is commonly assessed by the estimated glomerular filtration rate (eGFR), and calculated using formulas that incorporate endogenous markers like serum creatinine (SCr) and cystatin C. eGFR is a good estimate of overall kidney function and eGFR decline has association with CKD progression and mortality (1–3). Despite widespread use, eGFR alone does not provide information regarding injury to specific kidney cell-types (4, 5). Furthermore, a single GFR evaluation does not provide sufficient information about reversibility of current injury(6), and substantial alterations in serum creatinine and eGFR could also result from completely reversible hemodynamic changes(7). Urinary protein quantification, i.e. proteinuria, is also an orthogonal factor associating with the progression of CKD(8, 9). However, even together, these biomarkers often fail to comprehensively capture the extent and specific type of ongoing injury, somewhat restricting diagnostic and prognostic utility. Importantly, a significant amount of injury has to have accrued before increased serum creatinine is clinically detectable(10, 11).

The gold standard for diagnosing and monitoring kidney disease has therefore been a kidney biopsy(12). Biopsies provide understanding of compartment specific injury in the kidney (e.g. glomerular vs non-glomerular disease), understand ongoing pathologic processes as well as provide some information on disease progression (e.g. progression of fibrosis or IF/TA). To gain better pathogenetic understanding from biopsies, the field has layered on specific molecular diagnostic tools(13) to kidney biopsies. Single-cell RNA sequencing (scRNA-seq) from tissue is particularly promising, and can illuminate cell-type specific gene expression, and can profile how specific genes within cells mirror cellular injury and/or disease dynamics within kidney cell types(12, 14).

Although a valuable diagnostic tool, kidney biopsy has inherent limitations. First, there is sampling bias particularly relevant to diseases with patchy spatial involvement (e.g. FSGS, focal proliferative GNs or ANCA vasculitis) – a disadvantage that also spills over into biopsy based molecular diagnostics. Other issues include inter-observer variability and clinical risks such as bleeding complications, and arteriovenous fistula formation(15, 16). Additionally, biopsies are challenging in certain patient populations, including pregnant patients, those with bleeding diathesis, or a solitary kidney. These factors limit the performance of repeated biopsies and many times preempt biopsies altogether (12).

To fill this gap, a non-invasive method which has shown potential in research contexts, and could reflect compartment specific kidney pathology, is the quantification of urine cell pellet gene expression (17, 18). Single-cell data has demonstrated that expression of certain transcripts are indeed cell-specific(19). This raises the under-explored possibility that quantification of such genes in urine cell pellet could correlate with injury to their cells of origin(17). Importantly, cells in the urine reflect both kidneys from all filtering nephrons. Increased damage to specific kidney cells such as podocytes increases the urinary loss of these cells. For example, Wiggins et al. demonstrated that podocyte-specific markers can be detected in urine and correlating with podocytopenia in the biopsy(17, 20). Accelerated “podocyturia” correlated with increased podocyte loss in glomeruli in glomerular diseases but not in non-glomerular diseases, providing a non-invasive and semi-quantitative marker of both disease progression and cell-type specific injury(20, 21). Injured cells could increase transcription of injury-related genes, which could also be detected in urine when shed(22). In kidney transplantation, studies have demonstrated substantial overlap in gene expression between urinary cells and kidney allografts, with genes upregulated in rejection-vs non-rejection urine corresponding to kidney biopsy transcriptome changes (18, 22). The diagnostic utility of all these findings has not been fully explored in the context of native kidneys(18).

Taking advantage of this data, we combined recent information from single-cell data obtained in homeostasis and during injury, with the concept that cell-type specific genes would be detectable and quantifiable in the urine. We surmised that specific genes in the urinary cell pellet may hold cell- or compartment-specific information regarding ongoing kidney injury that can be assayed serially and in a non-invasive manner. We developed and validated a urine cell-pellet assay to quantify 10 genes to reflect specific aspects of kidney injury and demonstrate the utility of component genes in screening, correlation with pathology, and prognostication providing a non-invasive means for evaluating kidney disease (23). In this manuscript, we detail our assay development and report clinical findings from an unselected CKD cohort.

## Methods

### Cases and Controls-Controls

The control group consisted of healthy volunteers and living kidney donors at a pre-donation visit. Healthy volunteer controls were pooled samples derived from individuals with no evidence of kidney abnormalities or known clinical diagnoses of kidney disease. These healthy volunteers were sampled multiple times to generate pooled urine pellets. Kidney donor patients were screened prior to donation and met stringent criteria for donation and therefore assured of kidney health. Enrolled donors subsequently proceeded with successful donation, affirming their preserved renal function. Demographic details and laboratory findings were collected for these controls (if available). **Cases:** Patients were enrolled under an approved IRB protocol through the Yale Biorepository for Kidney Diseases which allowed to collect urine from participants (HIC# 2000027890; HIC# 2000036179). Participants from September 2023 to September 2024 were approached on the day of their scheduled outpatient kidney biopsy. This ensured that enrollees (cases) only included individuals with pathology significant enough to warrant a biopsy and a corresponding biopsy confirmation of diagnoses. Urine was used only if the specimen was obtained before their biopsy to avoid contamination of urine cell pellets with biopsy trauma-related entry of kidney parenchymal cells. The epidemiologic, clinical characteristics of cases, which consisted of participants with diverse/unselected renal biopsy diagnoses were collected.

### Patient Informed Consent/Data Collection

Prior to sample collection, informed consent was obtained from participants. Fresh urine samples and leftover urine samples obtained from the Yale Biorepository (HIC# 2000027890) were used to generate pellets. Each sample was assigned a unique study number connected to the biobank code. Clinical data for analysis were obtained from the biobank and electronic medical record and entered into a HIPAA-safe database without identifiers.

### Urine Collection

Urine samples were collected from unselected adult patients who were scheduled for a clinically indicated kidney biopsy (cases) and from healthy volunteers or donors (controls) as detailed above. The first samples provided by patients in the morning, before the biopsy procedure were collected to minimize any risk of biopsy-related injury altering the urine mRNA profile. Samples were collected from enrollees at the Yale Saint Raphael Campus over the 1-year study period.

### Volume of Urine for Assay

A specialized 220 ml EO sterile specimen container with lid (#76532-302, Avantor, USA) was used to collect the urine samples. A three-step layout for sample collection, processing, and mRNA quantification has already been published(24). This methodology specifically minimized RNA degradation during collection and transport, versus hospital-derived generic urine cups. About 10-15 mL of urine was collected in a 15 ml conical flask and simultaneously sent to Yale clinical labs for routine urinalysis, urine protein concentration, and urine creatinine measurement. The remaining urine was transferred to the research lab for further processing.

### Urine Transport

The urine samples were transported in thermally insulated bags containing reusable, leakproof refrigerant gel ice packs (THERMOSAFE, #420L78) pre-cooled to 4°C the day prior to use. The urine collection cups were positioned at the center of the ice packs within the insulated bags to ensure stable temperature control during transit. Upon arrival, the samples were stored at 4°C and were processed within 4 hours.

### Urine Processing

Total urine sample from a sterile cup was divided equally into 50 ml sterile centrifuge tubes with a sterile glass pipette and autodispencer. The sample was mixed to make sure that no fraction of cells remained stuck to the urine container. A maximum 45ml of urine was added to a sterile 50-ml plastic centrifuge tube. The centrifugation was done at 2000g at 4°C in a table-top centrifuge (Beckman-GS-6R) using swing-out rotor for 25 minutes. Two 2-ml aliquots and one 15 ml aliquot of urine supernatant were collected and stored at liquid nitrogen and −80°C, respectively, for future assays (proteomics, exosomal isolation/analyses). The remaining supernatant was discarded. The final urine cell pellet was resuspended in each 50 ml tube using 500 μl of cold diethylpyrocarbonate (DEPC)-treated PBS (pH 7.4), washed by gently pipetting with 1 ml sterile tips, and transferred to a new pre-cooled, labeled 1.7 ml Eppendorf tube (RNAse-free). To recover any pellet residue at the bottom of the 50 ml centrifuge tube, add another 500 μl of DEPC-PBS. After gently pipetting, transfer the contents to the same 1.7 ml tube. The resuspended pellet material in each 1.7 ml Eppendorf was then centrifuged at 13000 rpm for 5 minutes at 4°C in a mini-centrifuge (Eppendorf-5430 R), and the supernatant was discarded. Cell pellets were resuspended in 350-500 μl of RLT buffer containing β-mercaptoethanol (5 μl/ml), depending on the size of pellets. The samples were stored at −80°C for further use.

### RNA isolation from urine pellet

The stored urine pellets were removed from -80°C just before isolation and thawed by placing the samples on ice for 10 minutes. Following the manufacturer’s recommendations, RNA was isolated using the RNeasy Mini Kit (#74106; Qiagen). The overall yield of isolated RNA was low; hence, we optimized the kit protocol by performing the final elution in 30µL of RNA-free water to enhance the RNA concentration per microliter for cDNA preparation in the next step. To compare RNA isolation quality between our protocol and samples stored in RNAlater solution, we used the same Qiagen kit after giving an additional wash with RNAse-free water before the cell lysis step.

### RNA quality and quantity

RNA concentration was measured using NanoDrop-2000/2000c (Thermo Scientific, USA). The 260/280 ratio of RNA samples were quantified and used to determine samples not carried forward for reverse transcription. Reverse transcription was performed immediately after RNA isolation to prevent degradation due to freeze-thaw or batch-to-batch variation, using high-efficiency Superscript IV (Thermo Fisher), which has high sensitivity (≥10 pg template cDNA). Our first panel of three genes includes *18S rRNA* as an internal control to assess the quality of cDNA for subsequent analysis.

### Selection of mRNA Gene Panel

To develop a urine cell pellet mRNA assay for detecting kidney-compartment or pathogenesis-related genes, we strategically selected our gene candidates for the assay. We included genes for their (a) cell-specificity confirmed by kidney single-cell (Sc) RNAseq or single nuclear (Sn) RNAseq, (b) identifiable changes at the single cell level during injury, (c) their published correlation with pathologic changes in biopsies (d) confirmed cell-specific detection in Urine scRNAseq datasets. These criteria allowed for a high likelihood for detection and quantification in urine cell pellets by PCR. The gene candidates we selected based on these characteristics are a parsimonious representation of kidney pathology in urine. A brief description of each gene included in the Nephro-Dx panel is provided below.

- *<u>Podocyte-specific Genes</u>*: Podocin(*NPHS2*) gene is present in both fetal and adult kidneys, situated at the podocyte slit diaphragm(25–28), with minimal extra-podocyte sources of expression(28, 29). Nephrin (*NPHS1*) plays a critical role in the glomerular filtration barrier(30) and in the kidney, expression is restricted to podocytes(31). Urinary shedding of *NPHS2* & *NPHS1* has been associated with podocyte loss in experimental models(17) and has associated with prognosis in *<u>some proteinuric human</u> <u>diseases</u>*(21, 32).
- *<u>Proximal Tubular injury:</u> KIM-1*(*HAVCR2*) is an injury marker that is upregulated in proximal tubular cells(19, 33) - the critical cell type involved in acute tubular injury from diverse etiologies. The soluble form of human KIM-1 protein can be detected in the urine of patients with ATN(34).
- *<u>Collecting duct and Connecting tubule:</u>* Aquaporin-2 (*AQP2)* gene expression in the kidney is restricted to collecting duct principal cells and connecting tubule cells(35, 36). We hypothesized that urine *AQP2* may change in conditions of tubular atrophy or injury, similar to reductions in intra-parenchymal *AQP2*(28, 37).
- *<u>Fibrogenesis:</u>* Transforming growth factor-beta (*TGFB1*) is a central mediator of fibrogenesis and is upregulated in several forms of kidney injury, including CKD. Increased urinary TGFB1 protein levels have been correlated with tubulointerstitial fibrosis and glomerular sclerosis potentially due to enhanced renal macrophage infiltration and profibrotic signaling(38–40).
- *<u>Myeloid cell infiltration/Inflammation:</u>* Cluster of Differentiation 68*(CD68)* is a macrophage marker and is used to measure inflammatory cell infiltration in kidney disease. Elevated urinary *CD68* mRNA levels reflect active inflammation in conditions such as glomerulonephritis and interstitial nephritis(41, 42).
- *<u>Failed repair tubular cells:</u>* Vascular cell adhesion molecule-1 (*VCAM1*) is an adhesion molecule involved in leukocyte recruitment and plays a role in the repair response following kidney injury, particularly in the transition from acute kidney injury (AKI) to chronic kidney disease (CKD)(43).
- *<u>Progressive tubular injury:</u>* Shroom Family Member 3 (*SHROOM3*) Genome-wide association studies have identified genetic variants in the first intron of the SHROOM3 gene that are associated with CKD(44–46). In kidney transplantation, elevated *SHROOM3* levels at three months post-transplant correlated with increased allograft fibrosis and a decline in eGFR at 12 months(47). Increased tubulointerstitial *SHROOM3* also correlated with reduced GFR in native kidney CKD(47, 48). Our recent mechanistic data suggests increased tubular-specific SHROOM3 contributes to progressive CKD after injury via Rock-activation(49), and we hypothesized that tubular cell origin *SHROOM3* expression surveyable in urine is a likely signal of ongoing injury.
- *<u>Genes to reflect total cell content:</u>* 18S ribosomal RNA (*18s rRNA*) is the basic component of all eukaryotic cells and structural RNA for the small component of eukaryotic cytoplasmic ribosomes. This serves as a housekeeping gene, and widely used for normalization in quantitative PCR experiments due to its stable expression in various conditions and cell types(18, 22).
- *<u>Urinary epithelial shedding:</u>* Uroplakin 1A (*UPK1A*) is a specific marker of urothelial cells lining the urinary tract. Its expression in urine reflects cellular shedding from the urothelial lining(50–52), and potentially could serve as a control for cell-pellet cell and RNA content.
- *<u>Provisioning for panel expansion:</u>* We also surmised that such a qPCR panel could be sequentially augmented by addition of novel and robust gene candidates likely to be identified in subsequent studies, with minimal technical difficulty.

### Standards and plasmids

To develop a standardized method for quantifying our target genes in urine cell-pellet mRNA, we designed a control plasmid for each gene (i.e. as cDNA template) by using specific gene sequences from the gene’s mRNA sequence that could be detected by the corresponding TaqMan assay. For this, we identified the approximate location of each TaqMan assay primer (Thermo Fisher) on the corresponding mRNA sequence of the gene from NCBI-gene database. A sequence of approximately 150 base pairs in both 5’ and 3’ directions from the midpoint of each assay location was then designed for each gene. These unique control template sequences were synthesized and incorporated into vectors using the pUCIDT-AMP Golden Gate vector.

The gene vectors were then transformed into *E. coli* using One Shot™ TOP10, a chemically competent cell (C404003). After transformation, the cells were incubated overnight on LB agar plates containing ampicillin to select for transformed colonies. Once pure colonies were isolated, we performed Colony PCR to verify the presence of these genes in the transformed *E. coli* cells. The positive colonies were cultured overnight, and plasmids containing the desired genes were isolated using the Qiagen Maxi Prep Kit (catalog #12162). After obtaining the purified plasmid DNA, a master pool control template was prepared, containing 10^10 copies of each gene. For this, we calculated the molecular weight of each control plasmid to be able to estimate copy numbers per microgram or per unit volume.

### Assay Development

These plasmids insert sequences were first confirmed by NextGen sequencing. Next, in our laboratory, we also validated our in-house developed plasmids by qPCR. For this, we designed proprietary primers for each cDNA target sequence (Supplementary Table-1) and generated standard curves (10^1 to 10^9 plasmid copies at 1:10 sequential dilution) using SYBR Green to confirm the ability to produce accurate standard curves with these plasmids. Once the individual plasmids were validated by qPCR using SYBR-green, we proceeded to test each gene using TaqMan assays to ensure that the established standards performed as expected in the final assay. At this stage, we included urine cell pellet samples to ensure we could detect each gene in cDNA derived from urine pellets in a real-world scenario.

### Assay standardization

To standardize our assays, we used the plasmid controls of respective genes as templates to develop standard curves to quantify copy numbers of each gene per sample. This was done by using 10^1 to 10^9 copies of each gene as a template for generating a standard curve. Multiple standard curves were generated for each gene panel, with a minimum of three reproducible standard curves (R² >0.98) before finalizing each assay. A standard curve with at least five data points where theoretical copy numbers and obtained copy numbers were comparable in each run, and with an R² >0.98 was considered a good standard curve. Next, we ensured that CT values for a single sample or standard obtained from three different runs were within ± 1 SD when samples/ standard curve points run on different days were compared (day-to-day variation). When CT values were <2 SD for samples or standards, these occurred at very high concentrations (for standards) or at very low concentrations (<1000 copies for most genes). This allowed us to determine at least 5 data points on the standard curve which reflected the dynamic range of the assay and needed to be included in all plate runs. We also compared the raw copy numbers of all ten genes and calculated fold changes by generating a ratio using the average of pooled control samples. A comparative analysis was conducted to evaluate how well the expression of these genes correlated and how they can differentiate between control and pathology samples.

### Detection threshold

To establish detection thresholds for each gene in urine samples, qPCR of control cDNA was run simultaneously along with respective standards curve for each gene in every plate. False positives were excluded by running a negative control and a pool control with known copies in each plate to ensure consistency and minimize run-to-run variability. A master pooled control was generated by combining three pooled controls, and this pooled control was run on different days across different plates. Additionally, at least 10 pooled controls, collected at different times, were included in each plate. Absorbance ratios (260/280 nm) and RNA concentrations, as measured by Nanodrop, were plotted against the CT values of the ten genes to assess the correlation. Based on these data, we established thresholds for RNA purity and concentration. *18s rRNA* copies served as a housekeeping gene and were used for normalization in quantitative PCR. This gene is included in our panel-1 and we have used *18s rRNA* below 10^6 copies as a threshold to exclude patient samples for further analysis(18, 22). After calculating the copy numbers for all genes, we constructed a quality matrix based on the absence of amplification for one or more genes. We also considered whether the genes that failed to amplify were podocyte-related or non-podocyte genes, assigning a corresponding score to each case. We surmised that in cases of acceptable urine RNA yield, podocyte-specific mRNA could still be low if podocyte injury was less important in a given sample. These approaches enabled us to exclude samples with truly low RNA concentration or purity, and those cases where amplification failed for more than two non-podocyte genes-together reflecting unreliable urine pellet mRNA for analyses.

### Multiplexing

Once each gene’s TaqMan assay for detection/quantification and, each standard curve was validated, we developed three multiplex assays, each incorporating three genes from our panel, for the detection and quantification of the target urine biomarkers. For each panel, we customized 3 genes as multiplex TaqMan assays labeled with VIC, FAM, and NED probes. Multiplexing three genes in a panel was partly done for logistic reasons in the Applied Biosystems platform. Multiplexing also minimizes RNA requirements and plate-to-plate amplification variation, allowing simultaneous testing of three genes in a well for both patients and controls.

The three panels are as follows; Panel 1: *NPHS2, AQP2,* and *18S rRNA,* Panel 2: *NPHS1, TGFB1,* and *UPK1A* and Panel 3*: KIM-1, VCAM1,* and *CD68*. Another gene (*SHROOM3*) was included and tested using FAM-MGB-based singleplex TaqMan assays. Detailed information about the gene panel selection is already mentioned above in the gene selection paragraph, and probe IDs are provided in the Supplementary Table-2. All qPCR reactions were performed using StepOne instrument (Applied Biosciences, USA).

### Urinary gene quantification in cases/cohorts

All samples that passed the QC criteria were tested for ten mRNA signatures. TaqMan PCR was performed as mentioned in the methods section using customized TaqMan probes in three panels. In each well 5 ng cDNA was added, and samples were run in duplicate. For each test plate, pooled controls were also tested, and standard curves were included every time to determine the copy numbers per microliter.

### Pathology

Kidney biopsy reports, including immunofluorescence (IF) and electron microscopy (EM) findings, were retrieved from the Yale Renal Pathology database. Key biopsy parameters, such as the number of glomeruli, sclerosed glomeruli, interstitial fibrosis, tubular atrophy (IF/TA), and interstitial inflammation scores, were systematically recorded. Additionally, podocyte effacement and other relevant findings from the biopsy reports were noted and tabulated for subsequent clinicopathological comparisons. Locally reported pathology components (IF/TA, Tubular atrophy, injury) were tabulated from biopsy reports for histologic comparisons, and categorized as follows-none, mild, moderate, or severe as 0,1,2 or 3, respectively. *<u>Digital pathology</u>* was used to score the PAS-stained kidney biopsies, and quantitative scores were generated as previously described(53). A published automated digital pathology pipeline was used for quantitative digital pathology(53). A total of 10 scores were utilized for each biopsy which were related to glomerular (3/10), tubular (5/10), interstitial (1/10), inflammation-related (1/10) scores. These scores were numerically and continuously reported in a manner blinded to the operator. Digital pathologic scores were correlated with those reported by renal pathologists and with clinical outcomes to test their accuracy. Digital pathologic scores were used to compare against quantitative PCR data for ten genes to assess their specificity with compartment-specific injury on kidney biopsy (see below).

### Statistical analyses

*<u>Descriptive statistics</u>* (means and SD) were used to summarize the baseline clinical characteristics of Cases and Control cohorts and were compared using the chi-squared test and Fisher’s exact test. Normality of sample distributions was confirmed using Kolmogorov–Smirnov and Shapiro–Wilk tests. Univariate comparisons of continuous variables were done using unpaired *t* test (Mann–Whitney test for corresponding nonparametric analysis). *<u>Urinary mRNA data</u>* were reported both as copy numbers (from standard curve) and as fold changes (i.e. for each gene in a sample to average copy number of controls within the same plate) and analyzed using both parameters. To initially screen for associations between urinary mRNA and clinical/pathologic parameters, we used non-parametric Spearman correlations. For categorical comparisons between conditions, we used T-tests or ANOVA.

## RESULTS

### Controls

Urine samples from seven living kidney donors (Figure-1a). Kidney donors had completed screening and subsequently proceeded to successful donation signifying kidney health. Next, from 15 healthy volunteers, 60 urine samples were collected to generate 14 pooled control samples. Pooled controls enabled assessment of day-to-day variation in urinary mRNA levels and provided sufficient RNA yield to run the same controls across multiple qPCR plates. A total of 6 control samples had to be excluded due to absent pellets and/or pink deposits in the pellet (see methods & Supplementary Figure-1). Hence, qPCR analysis utilized a total of 19 control samples (Figure 1a). As expected, controls were younger on average, showed preserved renal function and normal laboratory findings, and lacked biopsy data, although some age overlap with cases was observed (Table-1).

**Figure 1:**
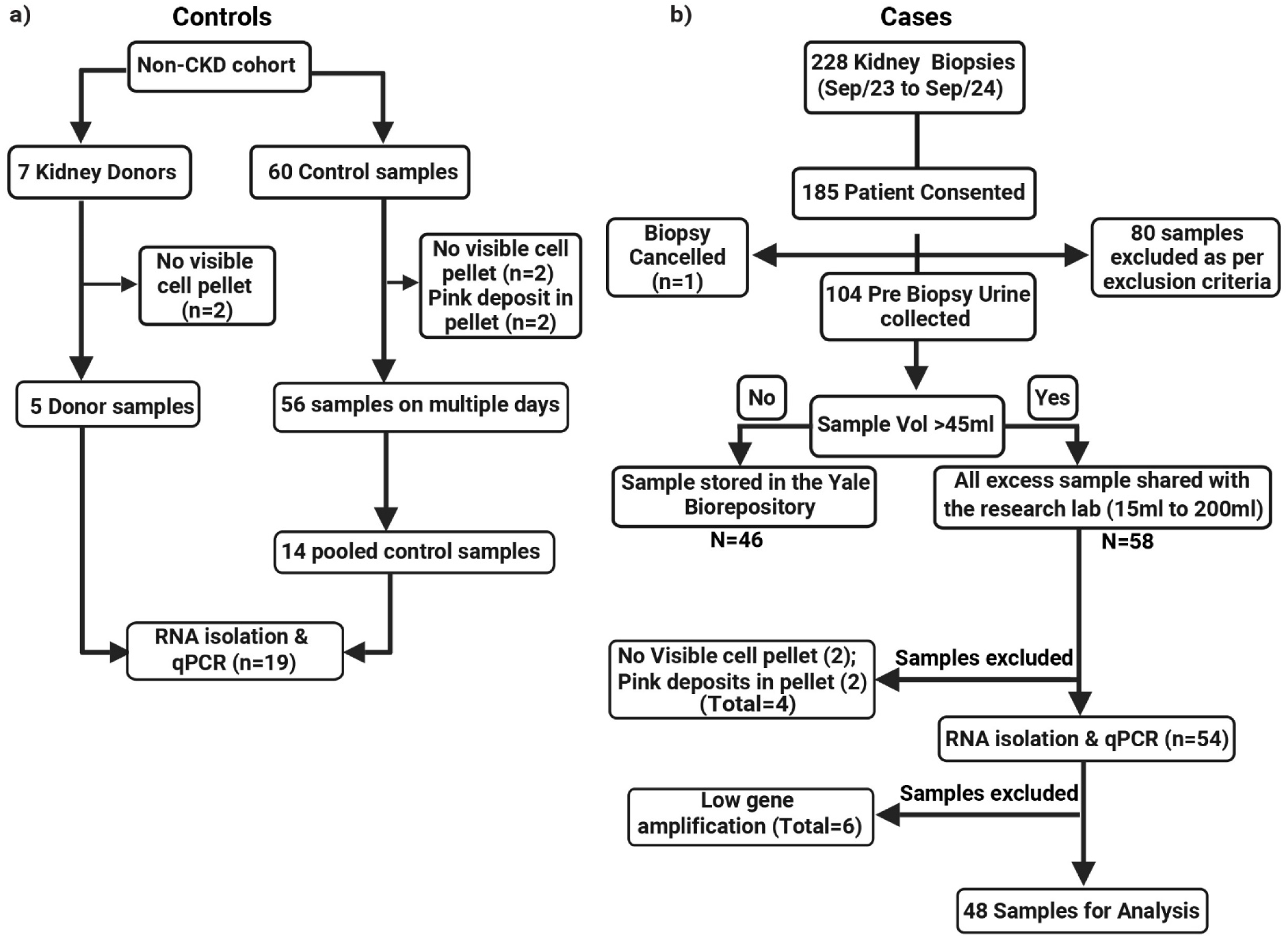
Overview of controls and cases used for urinary mRNA analysis. **a) Controls:** A total of 7 kidney donors provided urine samples, with 2 excluded due to no visible cell pellet, leaving 5 donor samples for RNA isolation and qPCR. From 15 healthy controls, 60 samples were collected, with 4 excluded due to no visible pellet and pink deposits in the pellet. From the remaining 56 samples collected on multiple days, 14 pooled control samples were used, and qPCR analysis was performed on 19 control samples. **b) Cases:** Between September 2023 and September 2024, 228 kidney biopsies were performed. Of these, 185 participants provided consent, and pre-biopsy urine samples were collected from 104 participants. Cases with urine sample ≤45 mL (N=46) were stored in the Yale biorepository. Samples with a total volume >45 mL (N=58) were collected and processed. After processing, 4 samples (2 with no visible cell pellet and 2 with pink deposits in the pellet) were excluded. RNA isolation and qPCR were done in 54 samples. All cases where total RNA was <100 ng, and the mean of *18s rRNA*, *TGFB1*, and *AQP2* were below controls, were also excluded. Finally, 48 cases were used for statistical analysis.

**Table-1:** Table showing clinical details of cases and controls in the present cohort.

| Variable | Controls N (%) [Range] | Cases N (%) [Range] | P value |
| --- | --- | --- | --- |
| Participants | 19 (100%) | 48 (100%) |  |
| Age Mean ± SD | 39.5 ± 14.6 [22 - 73] | 55 ± 16.82 [24 - 90] | **<0.01** |
| Gender (Female) | 10 (52%) | 26 (54.13%) | 0.79 |
| Race |  |  |  |
| White | 8 (42%) | 28 (58.3%) | 0.192 |
| AA | 3 (15%) | 7 (14.5%) | 0.999 |
| Others | 8 (42%) | 13 (27.1%) | 0.166 |
| Renal Function and Proteinuria |  |  |  |
| Serum Creatinine Mean ± SD | 0.87 ± 0.13 [1.0 - 0.73] | 2.12 ± 1.57 [0.49 - 7.75] | **0.017** |
| eGFR (CDK-EPI) Mean ± SD | 93.8 ± 5.26 [100 - 89] | 48.81 ± 31.27 [130 - 7] | **<0.01** |
| Urine Pt/Cr Mean ± SD | 0.058 ± 0.04 [0.01 - 0.11] | 3.35 ± 4.01 [0.04 - 21.21] | **<0.01** |
| Primary Biopsy Diagnosis |  |  |  |
| FSGS | No Biopsies | 12 (25%) |  |
| Diabetic Kidney Disease |  | 10 (20.8%) |  |
| Lupus Nephritis |  | 5 (10.4%) |  |
| IgA Nephropathy |  | 4 (8.3%) |  |
| Others |  | 17 (35%) |  |
| Presence of Acute Tubular Injury |  | 28 (58.3%) |  |
| Glomerular Disease |  | 45 (93.3%) |  |
| Non-glomerular Disease |  | 2 (4.2%) |  |
| Dipstick Proteinuria |  |  |  |
| Negative | 7 (36%) | 10 (20.3%) | 0.563 |
| (+), (++) | 0 | 15 (31.25%) | **<0.01** |
| (+++) | 0 | 23 (48%) | **<0.01** |

Demographic, renal function, biopsy diagnosis, and proteinuria data are presented for controls and cases. Values are expressed as N (%), mean ± SD [range]. Significant differences ( $P < 0.05$ ) are bold. Abbreviations: eGFR, estimated glomerular filtration rate (calculated using the Chronic Kidney Disease Epidemiology Collaboration [CKD-EPI] formula); Urine Pt/Cr, urine protein-to-creatinine ratio (mg/mg).

### Cases

Between September 2023 and September 2024, a total of 228 outpatient kidney biopsies were performed (Figure 1b). Of these, 185 patients (81%) consented to participate in the Yale biorepository. For the present study, we included only urine samples collected prior to the kidney biopsy to minimize alteration of urinary gene expression from biopsy-related direct injury or the presence of hematuria in the samples. Of the 104 participants who provided pre-biopsy urine samples, 54 urine samples of sufficient quantity were used for RNA extraction. Detailed reasons for lack of availability of urine among biobank consented patients for qPCR assay are in Fig 1B, and principally resulted from insufficient urine volume after biobanking. Following quality control, 6/54 samples subsequently failed quality control (QC; see below) were excluded leaving 48 patients for clinical analyses. The demographic and clinical details of these 48 patients are given in Table-1.

### Gene Markers

We developed a urine cell pellet mRNA assay to detect specific genes to reflect injury to cell types and kidney compartments (see methods and Figure-2). We included 10 genes *NPHS2, NPHS1, AQP2, KIM1, CD68, SHROOM3, VCAM1, TGFB1, UPK1A* and *18s rRNA.* The detailed rationale and selection criteria for each of these genes are described in the methods section and summarized here as Table-2. To support the development of our urinary mRNA panel, gene selection was based on their cell-type-specific expression within renal parenchymal cells in biopsy as demonstrated by recent single-cell and multiomic kidney atlases(54, 55) (https://susztaklab.com), which defined cell-specific expression patterns of each of these genes, as well as their dysregulation between healthy and diseased states. We also confirmed that these same genes were detectable in cell-specific patterns in urinary cell pellet single-cell RNA sequencing datasets from patients with kidney disease and healthy controls, using published datasets (see Figure-3 &(19, 28, 56)).

**Figure 2:**
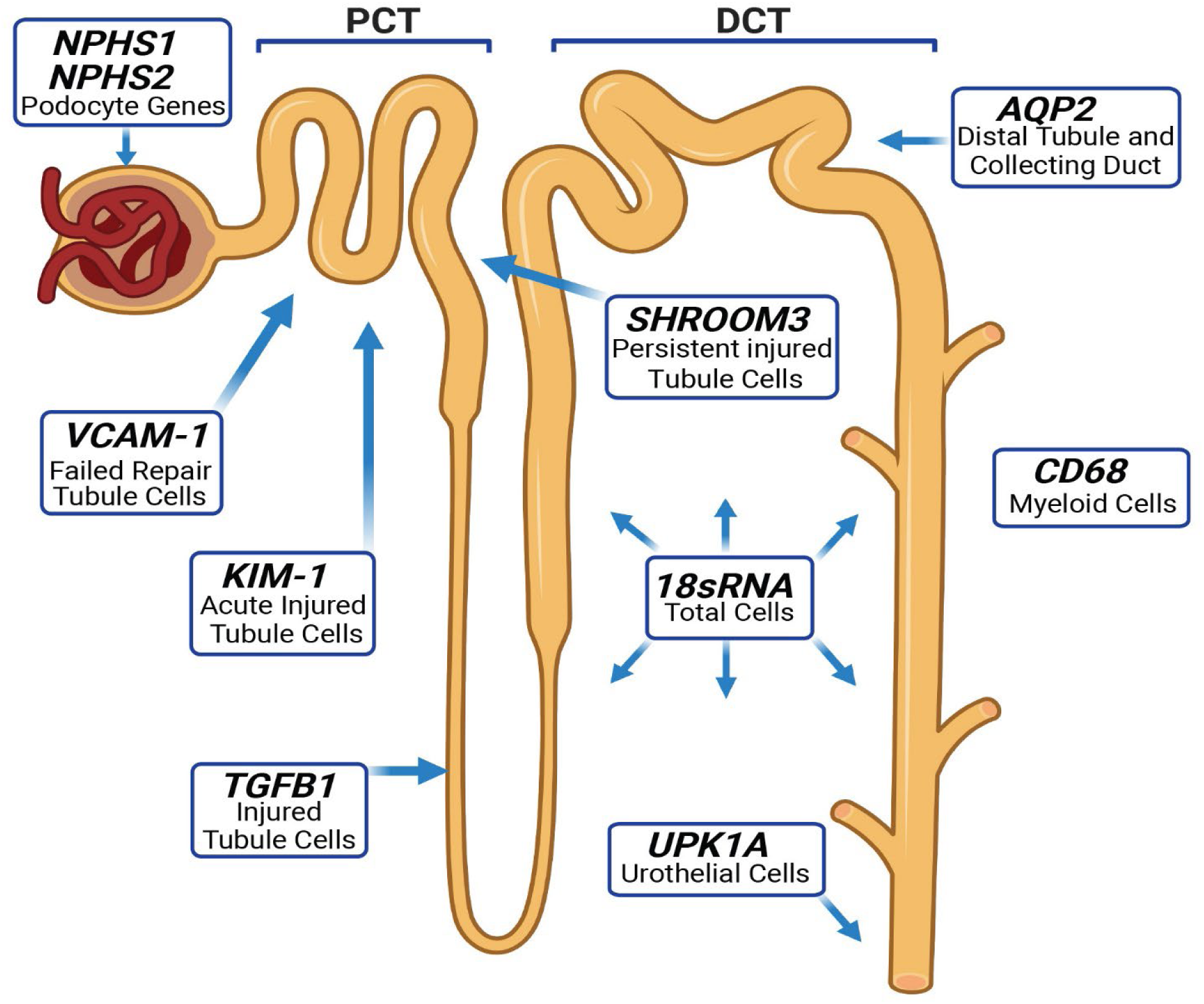
Overview of genes used in “Nephro-Dx” and their roles in specific nephron segments. *NPHS1, NPHS2* (Glomerular Segment): Specific to podocytes; urinary shedding reflects podocyte loss and disease prognosis. *KIM-1* (Proximal Tubular Cells): Marker of acute injury; elevated in tubular cells and detectable in urine. *AQP2* (Collecting duct principal cells and Connecting tubule cells): Urinary levels reflect tubular atrophy or injury. *TGFB1* (Tubulointerstitial): Central mediator of fibrosis; urinary levels correlate with tubulointerstitial fibrosis. *CD68* (Inflammation): Macrophage marker indicating inflammation in kidney disease. *VCAM1* (Tubular cells): Involved in AKI-to-CKD transition and tubular repair failure. *SHROOM3* (Tubular cells): Associated with persistent injury in proximal tubular cells, CKD progression, and fibrosis. *18s rRNA* (Total Cells): Housekeeping gene reflecting total cell content. *UPK1A* (Urothelial Cells): Urothelial cell marker; reflects shedding of cells from the urinary tract/bladder.

**Table-2:** Table showing significance and expression details of 10 genes used in Nephro-Dx.

| **Name** | **Cell of origin** | **Significantly changed with injury** | **Associated Pathologic changes** | **Detection in Urine ScRNA** | **References** |
| --- | --- | --- | --- | --- | --- |
| ***NPHS2*** | Podocyte | Increased | Podocyte loss, glomerular damage | Yes | (26) |
| ***NPHS1*** | Podocyte | Decreased | Podocyte loss, glomerular damage | Yes | (33) |
| ***KIM-1 (HAVCR1)*** | Proximal Tubule Cells | Increased | Acute Tubular Injury<br>Lymphoid inflammation | Yes | (19,34) |
| ***AQP2*** | Collecting duct and connecting tubule cells | Altered with tubular atrophy/injury (associated with AVP) | Water balance regulation | Yes | (26,38) |
| ***TGFB1*** | Tubulointerstitial cells, Macrophages, Injured tubular cells | Increased with progressive injury | Fibrogenesis | Yes | (41) |
| ***SHROOM3*** | Podocyte, Intercalated Cells, Distal Tubule, Proximal Tubule (injured). | Increased | Fibrogenesis | Yes | (47,46,48) |
| ***CD68*** | Macrophages and monocytes | Increased | Monocytic cells<br>Macrophages (circulating and resident) | Yes | (42,43) |
| ***VCAM1*** | Inflamed vascular endothelium<br>Failed repair tubular cells | Increased | Failed repair tubular cells | Yes | (44) |
| ***18s rRNA*** | All eukaryotic cells | Increased | Potential controls/QC |  | (22,18) |
| ***UPK1A*** | Urothelial cells | Increased | Potential controls/QC | Yes | (51,52) |

List of ten genes with cell of origin, injury-related expression changes, pathologic associations, and urinary detection validated by scRNA; references shown in the right column.

**Figure 3:**
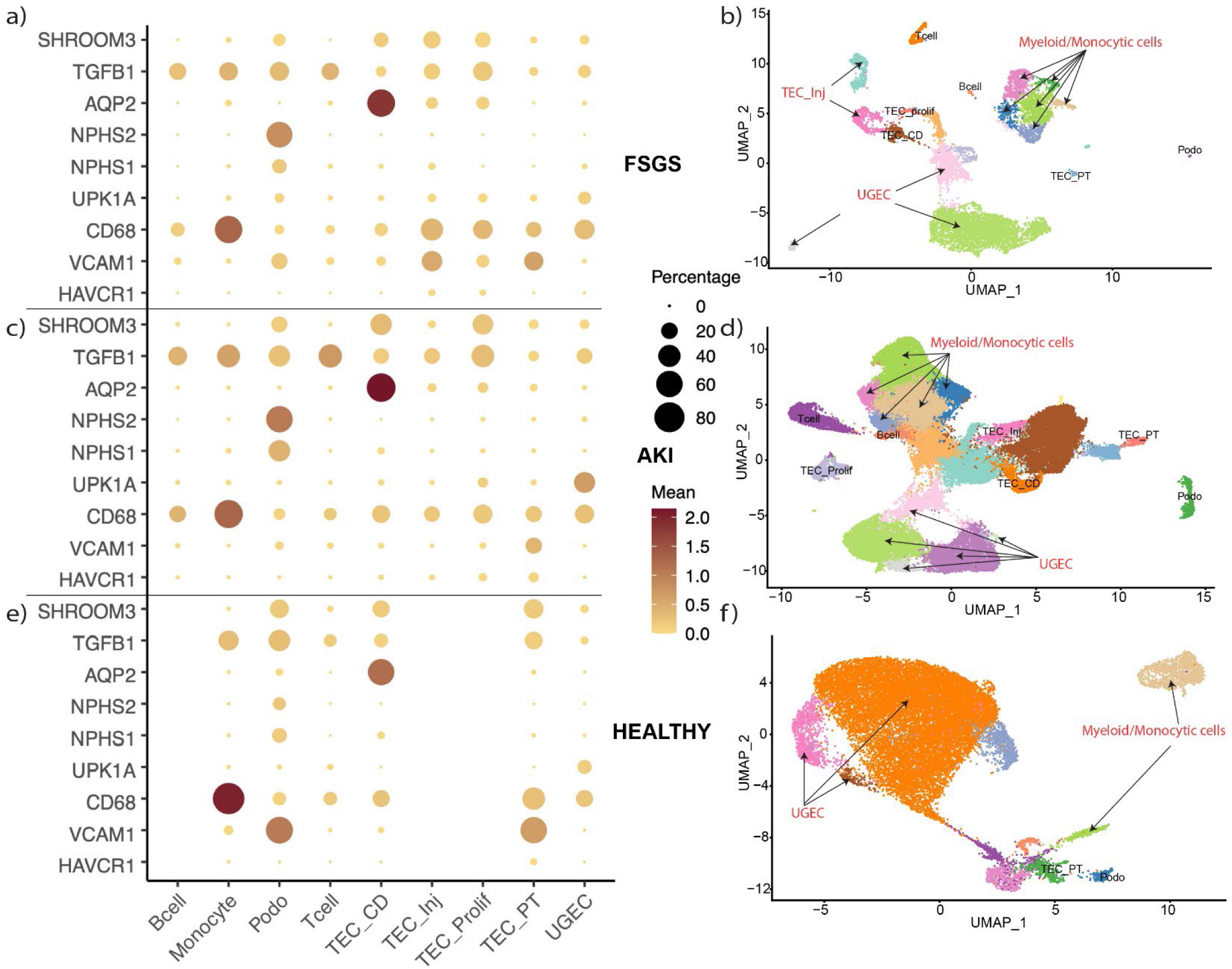
Single-cell sequencing data from urine samples obtained from diseased conditions or from healthy controls showing cell-specific and detectable expression of 9 of the 10 selected genes used in Nephro-Dx panel. Panels a, c & e are the dot plots from the datasets of FSGS, AKI and healthy controls, while b, d & f show the corresponding UMAP plots from these 3 different studies. Each color cluster represents a different cell type: B cell, Monocyte, Podo-Podocyte, T cell, TEC_CD-Tubular epithelial cell-Collecting Duct, TEC-inj-Tubular epithelial cell – Injury, TEC-prolif-Tubular epithelial cell – Proliferative, TEC-PT-Tubular epithelial cell – Proximal Tubule, UGEC-Urothelial Epithelial Cell.

### Assay development and technical validation

The details of urine collection, sample processing, RNA extraction, development of Standards, curve validation and multiplexing are provided under the respective sections in the methods. Briefly, a minimum of three runs were performed for each gene, and data was plotted as log-transformed copy numbers (theoretical versus calculated) corresponding to CT values at each dilution point (Supplementary Figure-2 & 3). Control samples were included during standard curve runs to determine detection threshold levels for each gene.

## QUALITY CONTROL METRICS FOR SAMPLES

### Urine pellet integrity in sample QC

We considered urine sampling, transport and processing as critical components and followed standardized operating procedures (see methods). The average volume of urine for controls and cases in the study was 392±185 ml and 40±28 ml respectively. We observed pink-colored pellets after centrifugation from 2 cases and 4 controls. These samples provided no RNA yield and had to be excluded from the analysis. These findings are consistent with previous reports, indicating crystals such as amorphous urate which can appear particularly in acidic urine as pink pellets(57–60).

### RNA Quantity and Purity in sample QC

The RNA concentration of the samples ranged from 3 to 180 ng/μl, with a mean ± SD of 27.89 ng/μl ± 30.57. An overview of the total RNA and purity matrix (260/280 ratios) in cases and controls are shown in the bar graphs in Supplementary Figure 4(a-b). We first compared the total RNA yield with the CT values obtained by qPCR for all ten genes (Supplementary Figure-5), and similarly compared purity ratios with CT values (Supplementary Figure-6). We observed that all cases with total RNA yield <100 ng had high CT numbers and low purity ratios and were excluded from the analysis. All excluded cases based on quantity and purity are highlighted as red dots in Supplementary Figure-5 & 6.

### Housekeeping Gene levels for sample QC

We included *18S rRNA* in Nephro-Dx as an internal control to assess the quality of cDNA obtained. The average copy number of *18S rRNA* ranged from 5 × 10^6 to 9 × 10^9 in both case and control samples. The detection of a sufficient quantity of *18S rRNA* copies in each sample provided confidence in RNA integrity and cell content of the sample, despite potentially lower copy numbers for other genes. Only 3 samples contained fewer than 10^6 copies of *18S rRNA*. Notably, these samples also exhibited low purity and low total RNA among the 6 excluded samples.

In summary, using these QC criteria, 6/54 cases and 6/67 controls urine that were collected in required volume were not usable for analyses.

## DIFFERENTIATING CASES VS CONTROLS

We first compared the abundance of each of ten genes between cases and controls. A standard curve and ∼10 control samples were included on each plate. We used the standard curve to obtain copy numbers for each gene, and then used these copy numbers to generate a fold change ratio for each sample over the mean copy numbers obtained from all controls in the same run (see Methods and Supplementary Figure-2 & 3). Thus, gene quantification was done in 48 patients and 19 controls, both by absolute copy numbers as well as by fold changes for each gene. Our analyses revealed significant increases in all 10 genes among cases vs controls. Summarized fold changes of each gene comparing cases vs controls are in Figure-4. Individual level data of copy numbers and corresponding fold changes for each of the ten genes are represented in Supplementary Figure-7 & 8, respectively. We also evaluated these values with or without normalization for urine volume (to represent gene abundance per ml of urine). In the latter analyses, differences between cases and controls were exaggerated, suggesting that cases (with kidney injury) had greater urine cell shedding per ml than controls (without parenchymal injury). Interestingly, fold changes of *18s rRNA* were also elevated in cases as compared to controls (albeit to a lower extent than other genes), raising the possibility that injured renal cells may express more *18s rRNA*. Normalization for urine creatinine concentration did not significantly alter the significant differences between cases and controls in Urine mRNA (not shown).

**Figure 4:**
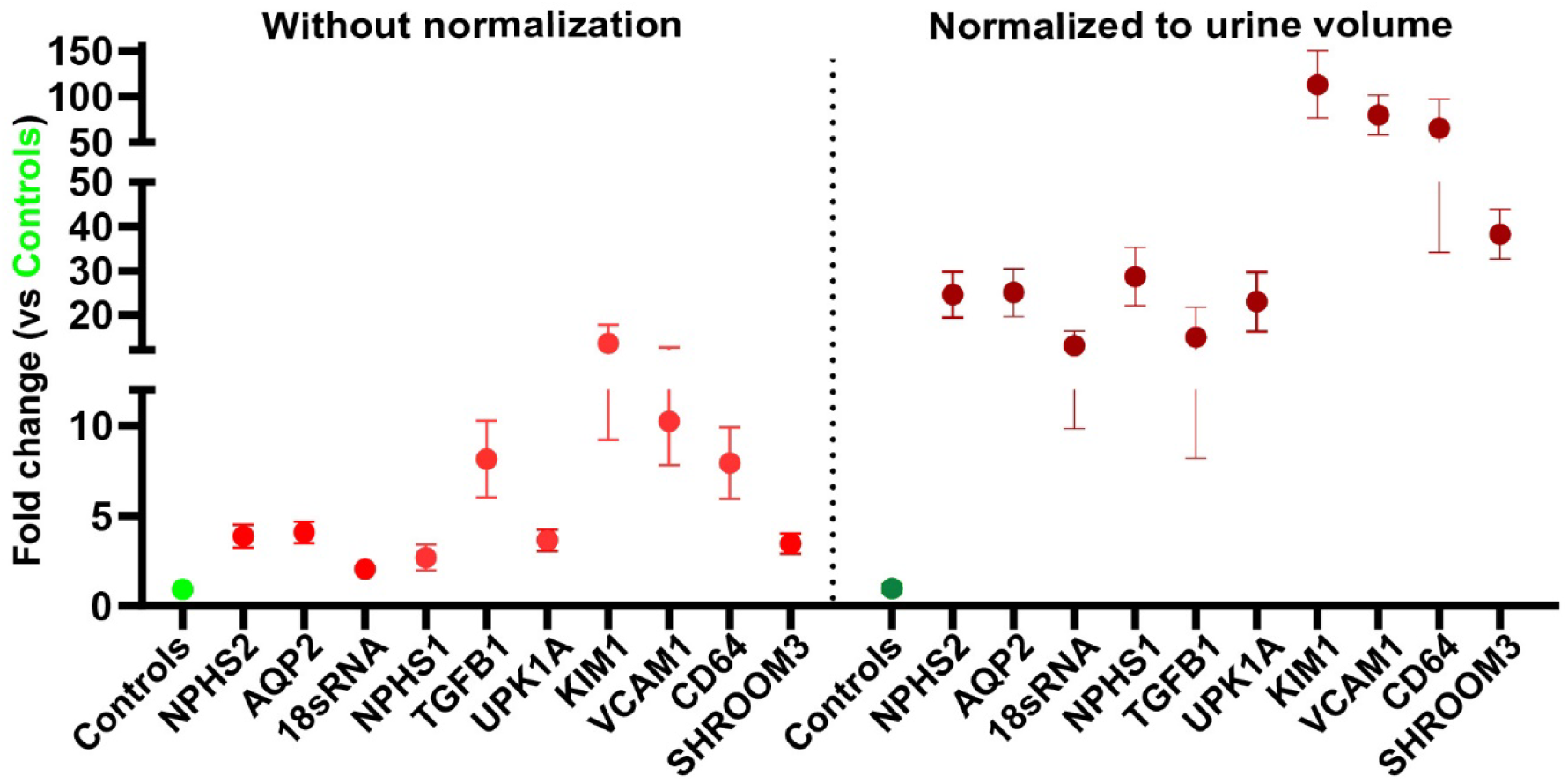
Urinary mRNA biomarker expression comparing fold changes in copy numbers between controls and cases with and without normalization to urine volume. Whisker plots showing mRNA fold changes normalized to controls for each of the 10 genes in cases vs controls, either without (left panel-light red dots) or with normalization to urine volume (right panel; dark red dots) for all 10 genes in control versus cases group (Dots/whiskers = mean ± SEM). The mean fold change for controls of all 10 genes are represented as a single dot (green or dark green color).

A logistic regression model for each gene demonstrated good discrimination between cases and controls, with all showing statistically significant associations (Table-3). These data reflected the potential of these biomarkers to differentiate any kidney injury vs healthy controls,

**Table-3:**
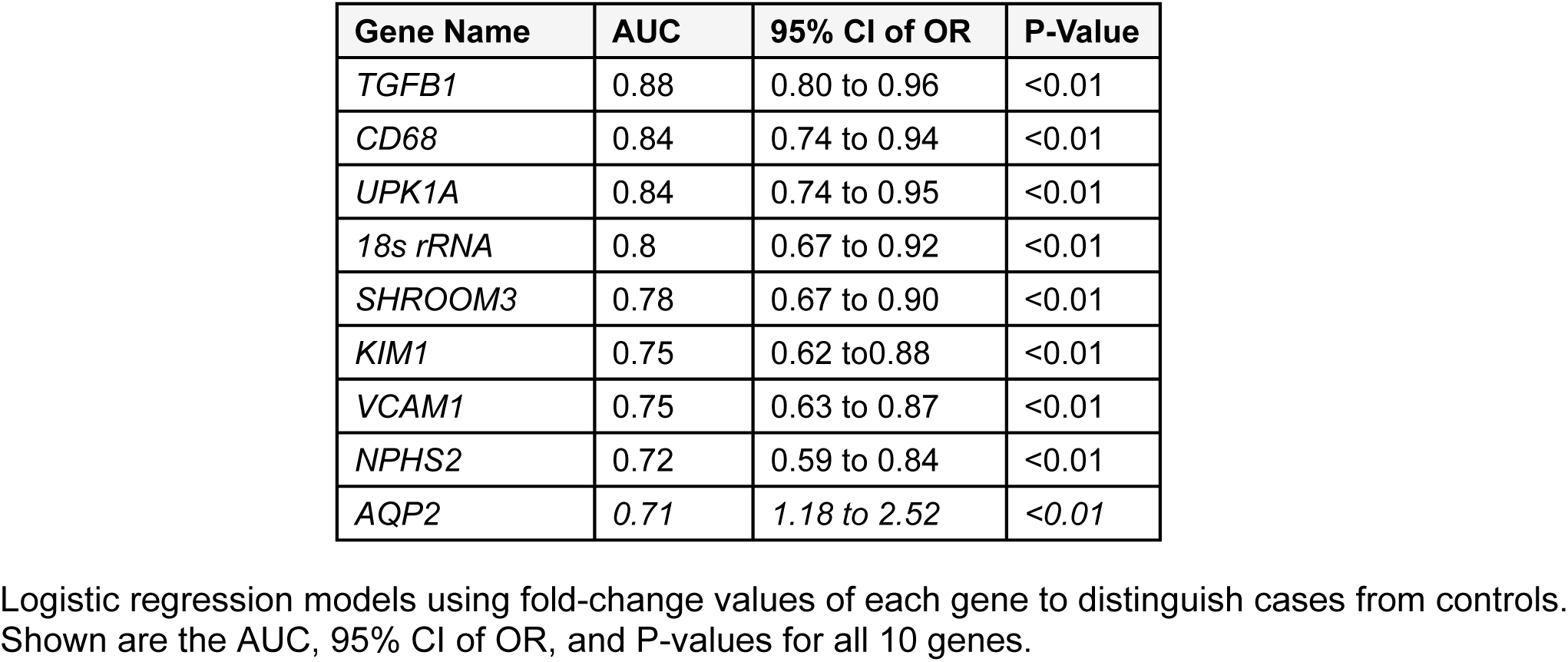
Table showing logistic regression models using fold change values for all genes to predict being in case or control group.

### Correlation of Urine mRNA with clinical characteristics

Since fold changes of cases-to-controls were highly correlated with copy numbers of each gene (Supplementary Figure-9 a-j), and were anchored by controls run in each plate with the samples (Supplementary Figure-9 k-t) and could best represent urinary gene excretion in the final assay, we used fold changes for most of downstream analyses.

#### eGFR-creatinine

We used the race-agnostic CKD-EPI equation to obtain eGFR-creatinine in our cohort. Importantly, none of the urinary mRNA showed any correlation with concomitant eGFR on the day of the biopsy, suggesting urinary shedding of cell pellets was not associated with glomerular filtration rate in this outpatient cohort (not shown).

#### Proteinuria

The copy numbers and fold-changes of *NPHS2*, *NPHS1*(podocyte-specific), *TGFB1* (fibrogenesis), and *CD68* exhibited strong correlations with the urine protein-to-creatinine ratio measured concurrently with renal biopsy (Table-4). These mRNA also correlated (except *TGFB1*) with dipstick proteinuria (Supplementary Table-3). Normalization for urine creatinine (to account for urine concentration) still retained significant correlations of urine *NPHS2* with urine protein: creatinine ratio [Spearman R=0.44; P<0.01; Supplementary Figure 10]. Additionally, *NPHS2*, *TGB1*, and *CD68* remained correlated with proteinuria levels two months after the biopsy (not shown).

**Table-4:** Clinical-Gene Correlations.

| Clinical data |  |  |  |  |
| --- | --- | --- | --- | --- |
|  | Gene Name | Correlation Coefficient | P-Value | Adjusted P-Value |
| Protein: Creatinine ratio<br>(at Biopsy in g/g) | *CD68* | 0.50 | <0.01 | 0.005 |
|  | *NPHS1* | 0.40 | <0.01 | 0.02 |
|  | *NPHS2* | 0.36 | <0.01 | 0.02 |
|  | *TGFB1* | 0.36 | <0.01 | 0.03 |

Spearman correlations of gene expression with protein: creatinine (g/g), showing correlation coefficients and significant values.

### Correlation of Urine mRNA with concurrent kidney biopsy features

Pathologic information is not well-reflected by conventional non-invasive tests of global dysfunction (creatinine or proteinuria). We hypothesized that our urine mRNA assay incorporating targeted genes would allow us to gain a non-invasive understanding of multiple facets of injury within kidney compartments and reflect specific pathology. We utilized our previously published automated digital pathology pipeline that annotates kidney compartments and quantifies normal/abnormal pathologic features (Supplementary Table-4) from slide-wide images(53) to examine the relationship between urine mRNA and various quantitative compartment-specific features obtained by digital pathology (Table-5).

#### Validation of digital pathology variables

We first evaluated if digital pathology features signifying compartment-specific pathology correlated with pathologist reports for the corresponding pathologic features (Supplementary Table-3). A manually quantified tubular area score (quantifying morphologically normal tubules using imageJ within each slide-wide image)(61) correlated linearly with normal tubular number, and inversely with interstitial fibrosis /atrophic tubule scores obtained by digital pathology. The tubular area score itself strongly correlated with kidney function (eGFR-creatinine), showing this was a readout for functioning kidney mass. A categorical IF/TA score (i.e. severity 0-3) obtained from pathology reports significantly correlated with interstitial fibrosis percentage in digital pathology. These data showed that quantitative digital pathologic features were representative of manual (imageJ) and pathologist reports from corresponding biopsies (Supplementary Table-3).

#### Correlation of Urine mRNA with Glomerular compartment pathologic features

A summary of biopsy diagnoses identified in our cohort are listed in Table-1. We evaluated podocyte-specific genes in our cohort with glomerular disease-related pathologic features. *NPHS1*-mRNA copies *<u>and/or</u>* fold changes correlated with normal glomerular number (normalized to area scanned per biopsy) (Pearson R= 0.47; P<0.01). On the other hand, urinary *NPHS2* correlated linearly with estimated glomerular area (glomerulomegaly), and quantile deviation (variation of glomerular area within a biopsy) (Table-5). These latter measures are known to represent ongoing glomerular injury and early glomerulosclerosis(49). The quantile deviation of the estimated glomerulus area also demonstrated associations with urine SHROOM*3* and *VCAM1* (Table-5).

**Table-5:** Correlation* of Digital Pathology Features with Gene Expression.

| Digital Pathology Features | Gene Name | Correlation Coefficient | P-Value | Adjusted P-Value |
| --- | --- | --- | --- | --- |
| **Glomerular** |  |  |  |  |
| Number of normal glomeruli | *NPHS1*<sup>#</sup> | 0.47 | <0.01 | 0.005 |
| Estimated mean glomerular area | *NPHS2* | 0.3 | 0.04 | 0.005 |
| Quantile deviation of mean glomerular area | *SHROOM3* | 0.55 | <0.01 | 0.02 |
|  | *NPHS2* | 0.46 | <0.01 | 0.005 |
|  | *VCAM1* | 0.45 | <0.01 | 0.005 |
| **Tubular** |  |  |  |  |
| Tubular Injury (None/Mild vs Moderate / Diffuse) | *KIM1*<sup>##</sup> | NA | <0.01 | 0.005 |
| Tubule slide-wide total area (Calculated Tubular Score) | *VCAM*<sup>#</sup> | -0.53 | 0.04 | 0.03 |
|  | *SHROOM3*<sup>#</sup> | -0.37 | 0.01 | 0.005 |
| Total number of atrophic tubules | *VCAM1* | 0.43 | <0.01 | 0.005 |
| % of atrophic tubules | *VCAM1* | 0.41 | <0.01 | 0.005 |
| Ratio of Atrophic: Normal Tubules | *VCAM1* | 0.54 | <0.01 | 0.005 |
| Number of normal tubules (based on pixels colors) | *VCAM* | -0.55 | <0.01 | 0.005 |
|  | *KIM1* | -0.42 | <0.01 | 0.005 |
| Severity of atrophy within tubules (based on pixels colors) | *SHROOM3*<sup>#</sup> | 0.3 | 0.04 | 0.005 |
| **Interstitial & Inflammatory** |  |  |  |  |
| % of abnormal interstitial area | *VCAM1* | 0.42 | 0 | 0.005 |
|  | *KIM1* | 0.36 | 0.01 | 0.005 |
|  | *SHROOM3* | 0.35 | 0.02 | 0.005 |
| % Percentage of lymphocyte | *KIM1* | 0.3 | 0.04 | 0.005 |

\*All digital pathology measurements were adjusted for Base\_Size\_1000000\_pw20 (see Supplementary Table-4). Associations used Spearman's correlation, with <sup>#</sup> for Pearson's (r) and <sup>##</sup> for Mann-Whitney U tests.

#### Correlation of Urine mRNA with tubular compartment pathologic features

We first evaluated the presence or absence of tubular injury on clinical pathology reports made by renal pathologists using a categorical scoring system (none-mild = 0; moderate to severe = 1). The tubular injury marker *KIM1* in the urine associated significantly with tubular injury score in biopsy (Table-5). The slide-wide total area of tubules identified (a surrogate for tubular cell mass and kidney function) were tested against tubular cell markers in our panel. Urinary markers representing injured tubules (*SHROOM3*, and *VCAM1*) were all significantly and negative correlated with tubular area from slide-wide images (Table-5).

Digital pathologic features of progressive tubular injury i.e. number and percentage of atrophic (dysfunctional tubules) in the biopsy, & ratio of atrophic: normal tubules on biopsy (based on distinct pattern of pixelation of atrophic features) strongly correlated with urinary *VCAM1* levels showing the importance of this as a marker of ongoing progressive injury(43). *VCAM1* & *KIM1* also correlated negatively with normal tubular pixelation (annotated as normal by digital pathology and normalized for biopsy area). Severity of atrophy of tubules (averaged over each biopsy) correlated with urinary *SHROOM3* – another marker of injury/ progression that we have reported(62) (Table-5). Hence, the combination of markers included in our panel provided strong correlations with both acute and/or progressive tubular injury on the biopsy.

#### Correlation of Urine mRNA with interstitial compartment pathologic features

Interstitial fibrosis is an estimate of accrued injury over time to the kidney and offers prognostic insight in all kidney diseases. Percentage of abnormal interstitium quantified by digital pathology correlated with interstitial fibrosis reported by expert pathologists (above), and strongly correlated with urinary *VCAM1* levels (Table-5), and modestly with *KIM1* levels and *SHROOM3*). *KIM1* is expressed in tubular cells but also transcribed by infiltrating lymphocytes (also known as *TIM1*)(63). The infiltration of lymphocytes into kidney interstitium is considered abnormal (quantified by digital pathology) and was associated with increased urinary *KIM1*(Table-5). Hence, our urinary marker panel provided key information mirroring abnormalities in the kidney interstitium.

### Prediction of kidney function decline using Urine mRNA

A central question in any kidney disease with or without a simultaneous biopsy is the risk of progression and later development of end-stage kidney disease. To test if our urine panel predicts subsequent progression, we followed patients in our biopsy cohort for ∼6 months from date of biopsy (n=47/48), and tested if urinary mRNA markers could predict disease progression. Patients whose eGFR declined > 20% from biopsy (n=16; 34%) were compared with those whose GFRs remained the same or improved (n=31; 65% Table-6). In this comparison, *NPHS2, AQP2, KIM1, CD68, SHROOM3 and VCAM1* were significantly elevated in the 20% eGFR decline group (Figure-5 a-f). The same result was observed when the outcome was considered any eGFR decline in 6 months (Figure-5 g-l). Hence, genes included in our urine mRNA panel which have established relevance to progressive injury (*VCAM1* and *SHROOM3*) also correlated with subsequent decline in renal function.

**Table-6.** Clinical characteristics by 6-month eGFR outcome: ≥20% decline vs stable/improved.

| Variable | no eGFR decline N (%) [Range] | ≥ 20% eGFR Decline N (%) [Range] | P value |
| --- | --- | --- | --- |
| Participants | 31 (65.9) | 16 (34.0) |  |
| Age Mean ± SD | 54.64 (± 16.48) | 55.56 (± 18.34) | 0.86 |
| Gender (Female) | 17 (54.8) | 8 (50) | 0.76 |
| Race |  |  |  |
| White | 18 (58) | 9 (56.2) |  |
| AA | 5 (16.1) | 2 (12.5) |  |
| Others | 8 (25) | 5 (31) |  |
| **Renal Function and Proteinuria** |  |  |  |
| Serum Creatinine Mean ± SD | 1.69 ± 1.05 [0.49- 4.20] | 3.03 ± 2.09 [0.68 -3.95] | **0.01** |
| eGFR (CKD-EPI) Mean ± SD | 55.57 ± 332.28 [130-12] | 36.45 ± 26.0 [103-7] | 0.06 |
| Urine Pt/Cr Mean ± SD | 2.66 ± 4.02 [0.04 - 21.1] | 4.88 ± 3.88 [0.2 -12.64] | **0.01** |
| **Primary Biopsy Diagnosis** |  |  |  |
| FSGS | 9 (29) | 3 (18) |  |
| Diabetic Kidney Disease | 5 (16.1) | 5 (31.2) |  |
| Lupus Nephritis | 3 (9.6) | 2 (12.5) |  |
| IgA Nephropathy | 3 (9.6) | 1 (6.25) |  |
| Others | 11 (35) | 5 (31.2) |  |

Demographic, renal function, biopsy diagnosis, and proteinuria data for cases (≥20% decline vs no decline [stable/improved] within 6 months). Values are reported as n (%) or mean ± SD [range]. Statistically significant differences (P<0.05) are shown in bold. Abbreviations: eGFR, estimated glomerular filtration rate (CKD-EPI equation); Urine Pt/Cr (UPCR), urine protein-to-creatinine ratio (mg/mg).

**Figure 5:**
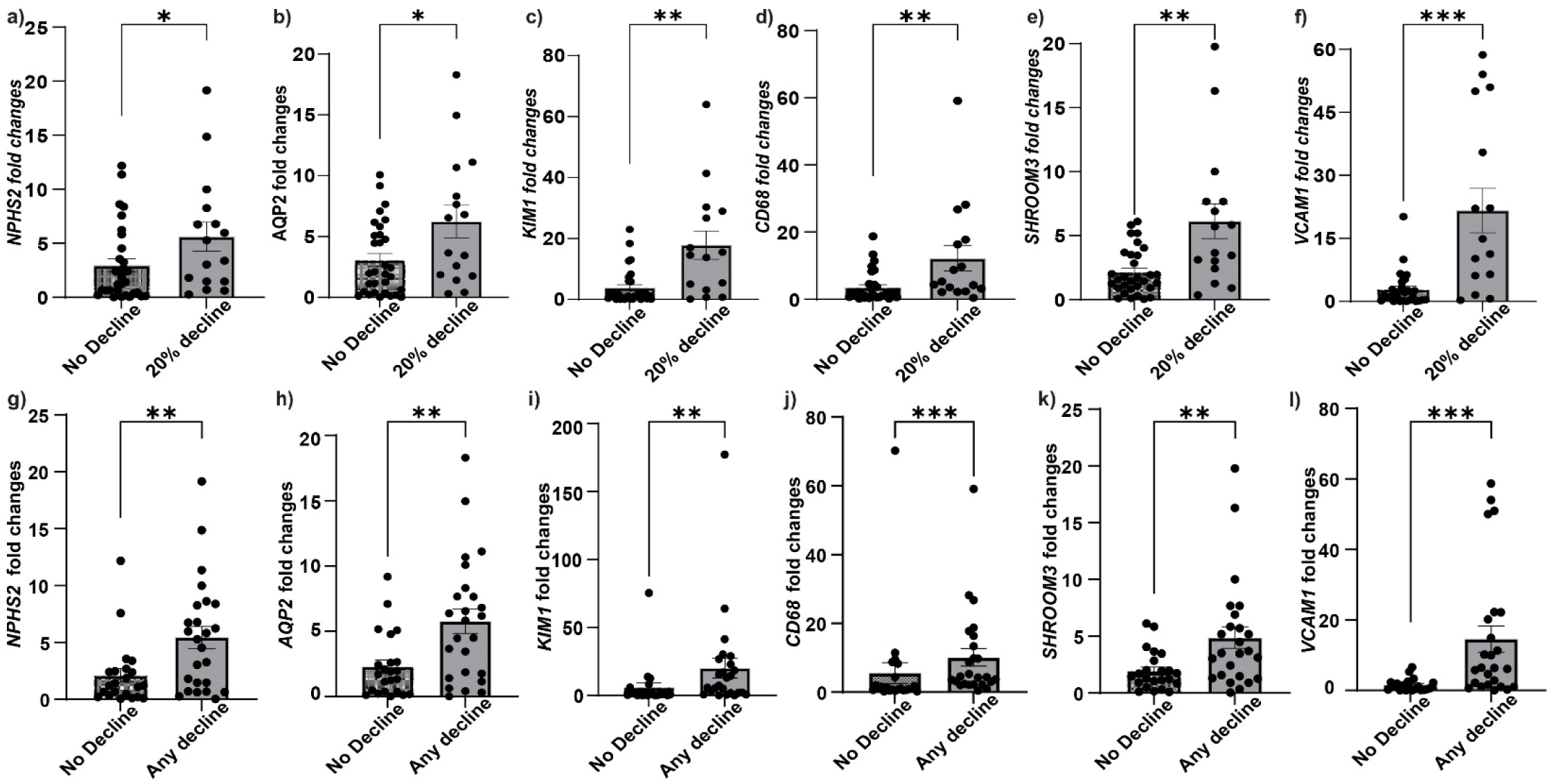
Urinary mRNA fold changes correlate with eGFR decline at 6 months. **(a–f)** Scatter plots compare urinary mRNA fold changes for 6 genes in patients with/without ≥20% eGFRdecline at 6 months: (a) *NPHS2*, (b) *AQP2*, (c) *KIM1*, (d) *CD68*, (e) *SHROOM3*, and (f) *VCAM1*. **(g–l)** Scatter plots compare urinary mRNA fold changes for 6 genes in patients with/without any eGFR decline at 6 months: (g) *NPHS2*, (h) *AQP2*, (i) *KIM1*, (j) *CD68*, (k) *SHROOM3*, and (l) *VCAM.* All six genes exhibited significantly higher urinary mRNA expressions in the decliner group. Statistical significance is indicated as *p*<0.05, \**p*<0.01, \*\**p*<0.001, \*\*\**p*<0.0001. [eGFR=estimated glomerular filtration rate].

## DISCUSSION

In this work, we develop a multiplex qPCR-based assay to reliably quantify 10 mRNA transcripts in the urinary cell pellet (i.e. Nephro-Dx) from a real-world cohort of patients with varying kidney pathology. These genes were selected strategically based on the specificity of expression, association with cell-type-specific injury, detection in the urinary cell pellet mRNA, and their pathogenetic roles in kidney compartment-specific injury processes. We report here that combinations of these carefully selected genes could not only provide cross-sectional/diagnostic information but also provide information regarding subsequent prognosis (progression of disease) and therapeutic responsiveness, filling key knowledge and product gaps in the field.

Our non-invasive assay has several advantages. First, this assay can be repeated serially and non-invasively at multiple time points and provide independent information to both *<u>invasive</u>* kidney biopsies (a specific test that cannot be frequently repeated), and *<u>existing non-invasive tests</u>* (creatinine and urinary protein, which are not designed to interrogate specific pathology or types of injury). Next, the genes incorporated here reflect specific aspects of injury, providing a multidimensional picture of the concurrent kidney disease. For instance, podocyte-specific gene quantification, reflected glomerular physiology (*NPHS1*) and glomerular pathology (*NPHS2*). Tubular injury/atrophy was accurately reflected by *KIM1* and *VCAM1*, while progressive interstitial replacement by fibrogenesis (a consequence of injury) was reflected accurately by *VCAM1*, and *KIM1* correlated with lymphocytic infiltration or inflammation in the biopsy. Notably, we tested the associations of our urine mRNA panel with pathology using a highly quantitative, automated, pathologic scoring system that is free from inter-observer bias and can be reliably repeated at any center regardless of the pathologist, demonstrating the robustness of our assay.

One of the genes in our panel (Shroom3) was selected based on the association of polymorphisms in Shroom3 with CKD in the general population(64, 65). Shroom3 mutations are highly prevalent in nearly 40% of the population, and associated with an increased risk of CKD. The increased risk of CKD occurs as these intronic mutations work to increase the expression of Shroom3(47). Our recent mechanistic work using unique animal models has shown its importance in ongoing tubular injury and fibrogenesis(49). While Shroom3 SNPs can be identified by genetic testing, every individual with Shroom3 SNPs does not progresses to CKD or dialysis, and our assay will provide an additional layer to risk-stratify such individuals, specifically at risk of progression from ongoing excess Shroom3. By incorporating biomarkers like *VCAM1*, *SHROOM3, NPHS2,* and *TGFB1* into clinical practice, clinicians could risk-stratify CKD patients, and potentially intervene earlier specifically in patients at high risk of progression. This could lead to better management strategies, whether by adjusting current therapies or introducing new treatments designed to slow disease progression, ultimately preventing or delaying the onset of ESKD. We also anticipate that combinatorial logistic regression models of one or more of the 10 genes, would be expected to provide information regarding both histologic correlates of injury as well as associations with subsequent outcome.

We paid specific emphasis to the technical robustness of our assay during every step of its development (detailed in the methods section) to provide a repeatable, quantifiable readout with acceptable day-to-day or plate-to-plate variation. The choice of the qPCR platform for our assay is also strategic to minimize cost and widen applicability to low-resource situations. Our assay also allows for customizing the panel of genes to a given situation-proteinuric glomerular disease or FSGS vs non-glomerular diseases. The platform of our panel can be flexibly expanded to include specific markers of distinct pathology-for instance, T-cell or B-cell infiltration, complement activation or other markers that may emerge in future research work. Hence, our panel lays the groundwork for future investigations into novel and additional markers that could further refine predictive models or serve as therapeutic response follow-up targets. The integration of molecular and clinical data in prediction models could ultimately catalyze more personalized approaches to kidney disease management in the future.

In conclusion, our findings using our panel could improve how kidney disease progression is monitored by offering a non-invasive liquid biopsy that signifies specific and concurrent pathology, integrating urinary mRNA biomarkers with clinical and histological data holds the potential to significantly improve patient outcomes, guide early interventions, and enhance the precision of personalized treatment strategies in kidney disease.

## Acknowledgment

MCM acknowledges funding from the NIH (grants R01DK122164, R01DK132274, and R21AI178705), and the Department of Defense, grants HT94252310454 and HT94252310441. MCM also acknowledges research support from the Blavatnik Fund at Yale (Accelerator award), and a pilot award from CTSA Grant UL1 TR001863, and Yale Centre for Clinical Investigation (Pilot Translational Award)

## Author Contributions Statement

A.K., GB and ZY contributed to the experimental work, data analysis, data interpretation, drafting, and editing of this manuscript; Z.S. was involved in the RNA sequencing data interpretation and data collation from published scRNAseq datasets; AR., E.M.T. H.S. and J.P. were involved in data curation, and editing this manuscript; S.P. provided interpreting pathological data from patients; M.S., C.K., D.J., and P.L. were involved in sample/data collection. D.J., and K.M. contributed to data curation and interpretation and editing of the manuscript; R.L. J.T contributed to data generation, perfoming the kidney biopsies; J.C.H. Data interpretation edited and reviewed this manuscript; D.M. data generation and interpretation. P.W. Data analysis and data interpretation, W.Z. Data interpretation edited and reviewed this manuscript. M.C.M. Conceptualized and designed this study, data interpretation, drafted/edited and supervised this manuscript. All authors reviewed and approved the manuscript.

## Data Availability Statement

The authors declare that all other relevant data supporting the findings of this study are available in this article and its Supplementary Information files. Additional data requests can be sent to Dr. Madhav Menon, corresponding author.

---

**Supplementary Figure-1:**
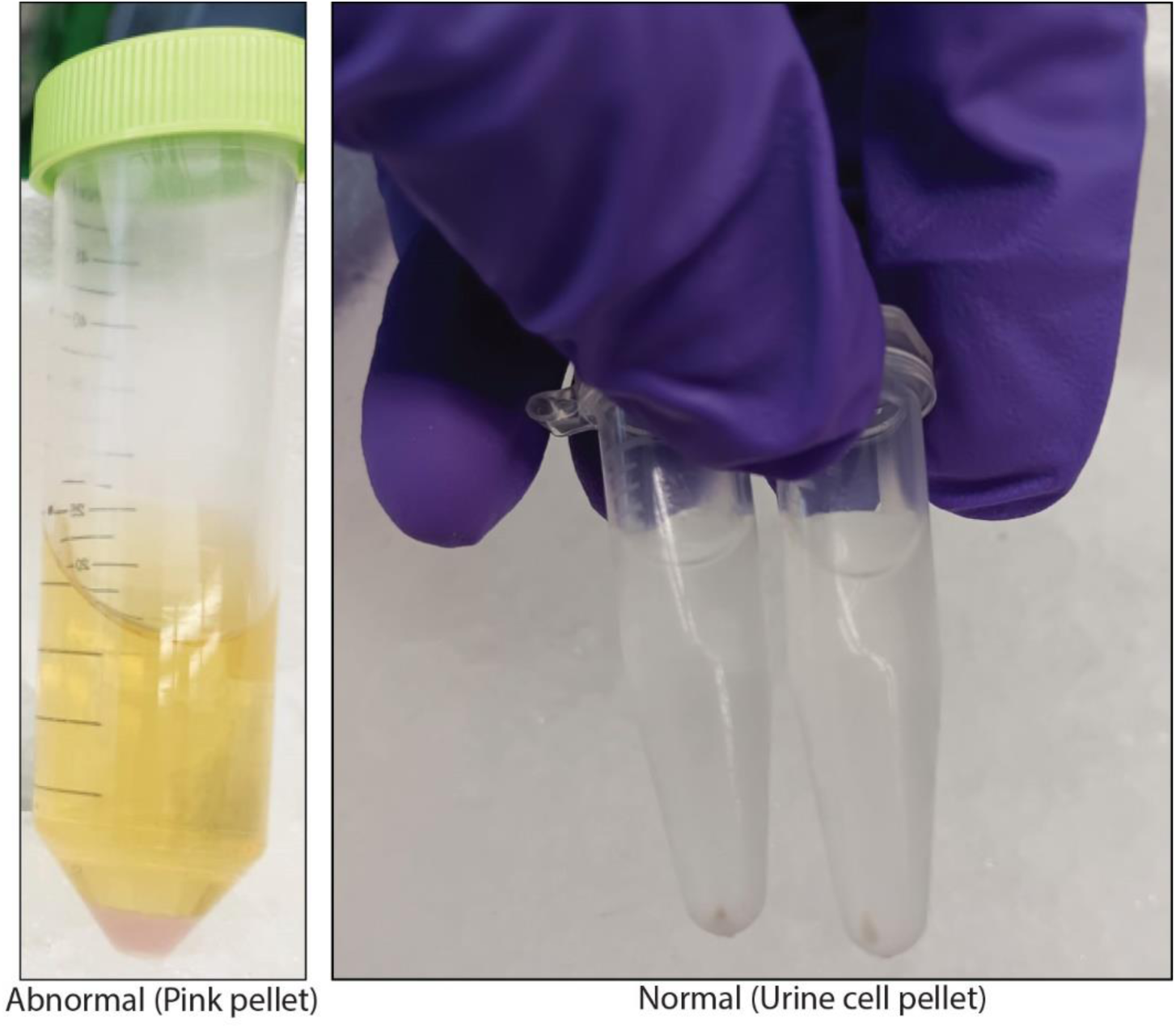
Comparison of urine cell pellets. The image on the left shows an abnormal sample with a pink pellet obtained after centrifugation. All samples with such pellets were excluded from the analysis. The image on the right shows normal urine cell pellets suitable for further processing.

**Supplementary Figure 2:**
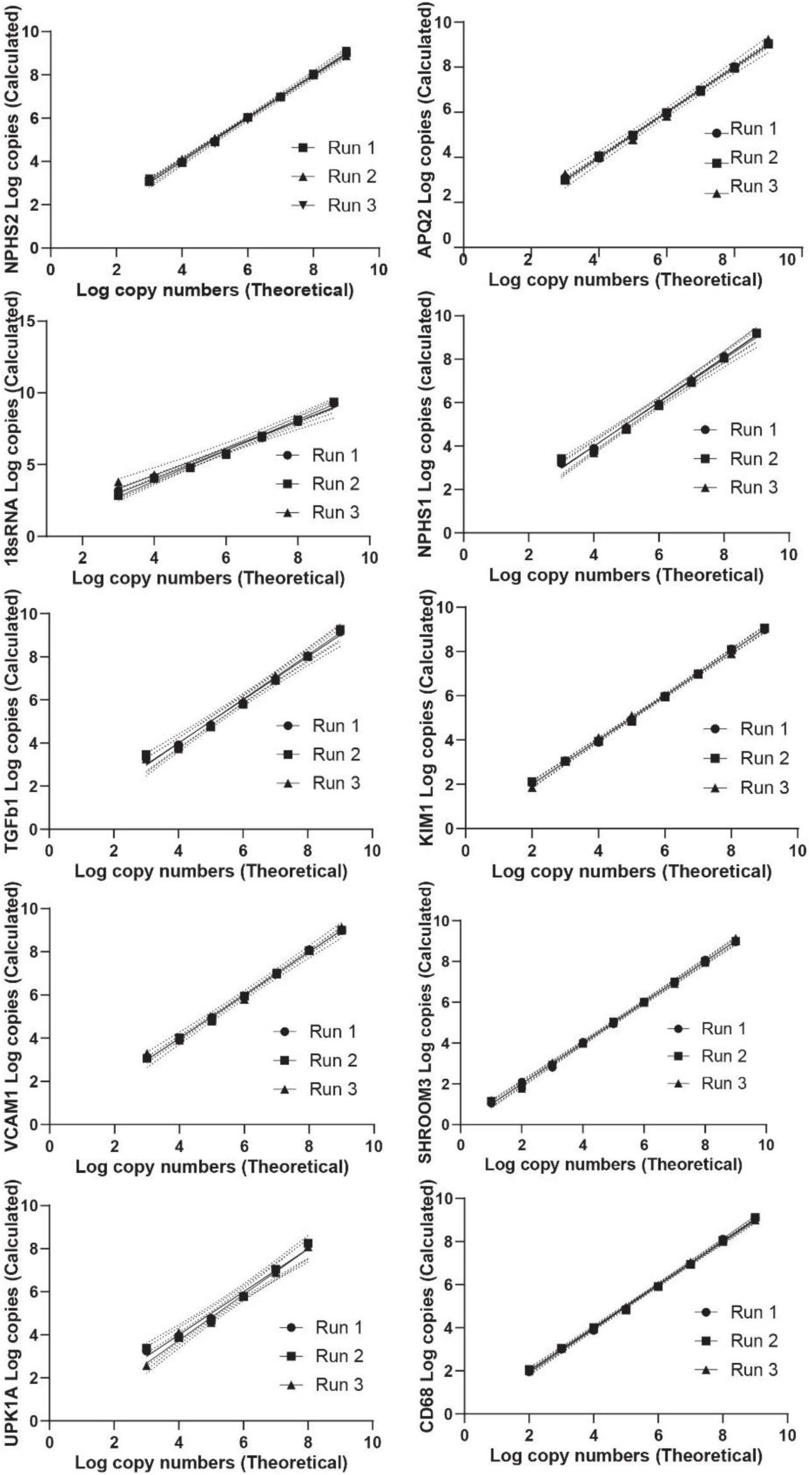
The figure panel presents linear regression plots for the standard curves of all 10 genes. A minimum of three standard curve runs were conducted for each gene, with data plotted as log-transformed copy numbers (theoretical versus calculated) at each dilution point.

**Supplementary Figure 3:**
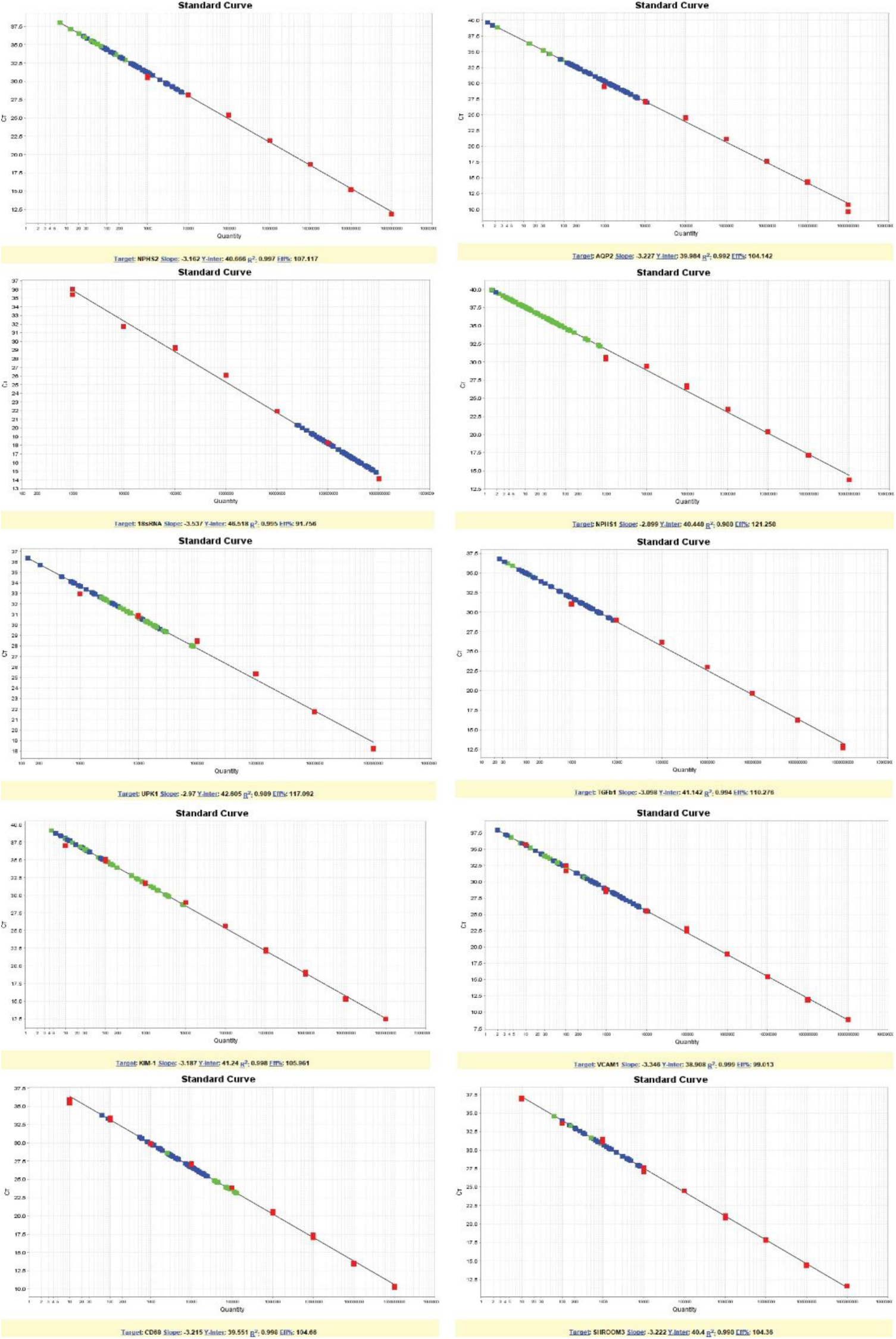
The figure panel presents software-generated representative standard curve graphs for all 10 genes. Each run achieved R² values >0.98, with at least five data points included for calculating copy numbers and interpreting the plate run. For each gene, a standard containing 10^10 copies was subjected to a 10-fold dilution, and qPCR was performed to generate the corresponding CT values.

**Supplementary Figure 4:**
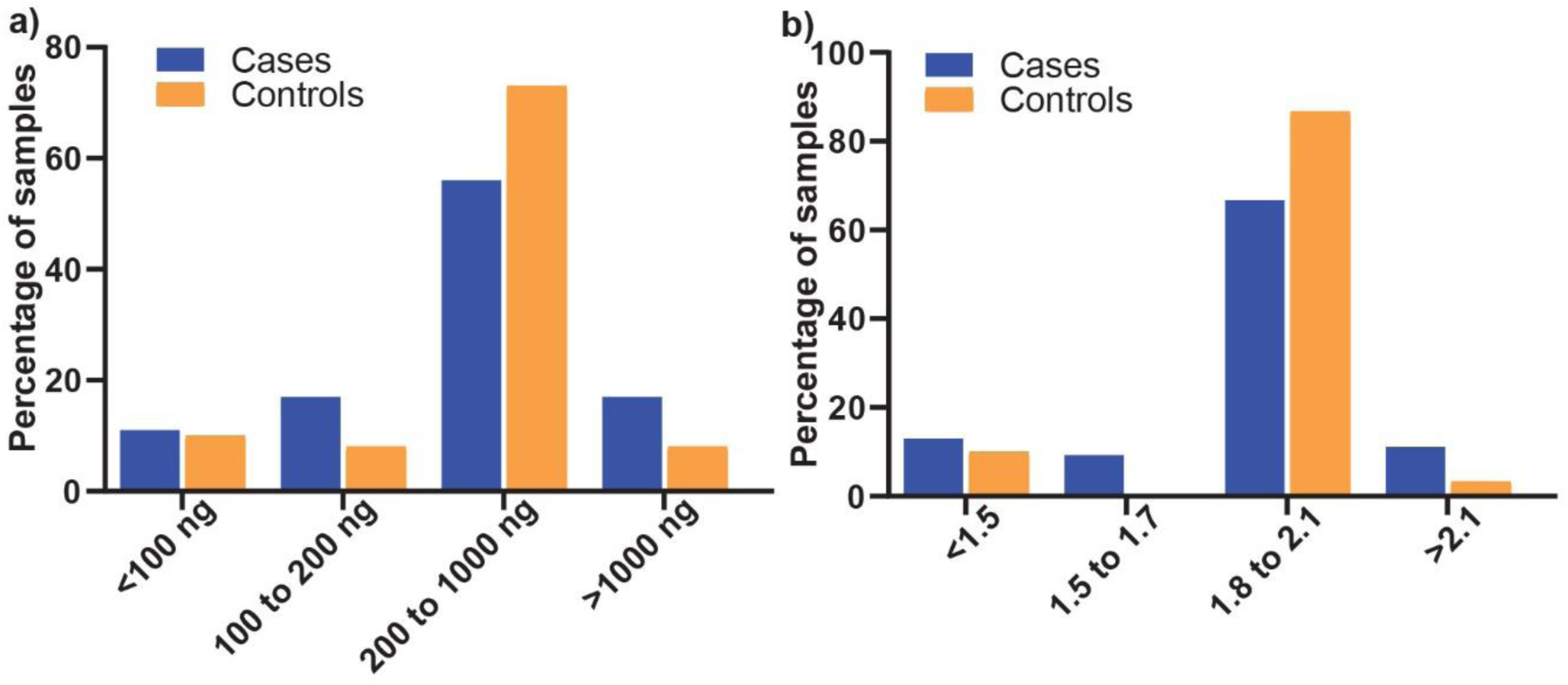
(a) A comparative bar-graph panel illustrating the total RNA isolated from the urine pellet in cases and controls. The samples where the total RNA was <100 ng were excluded from the analysis. (b) A bar-graph showing RNA purity (260/280 ratio) in the cases and controls.

**Supplementary Figure 5:**
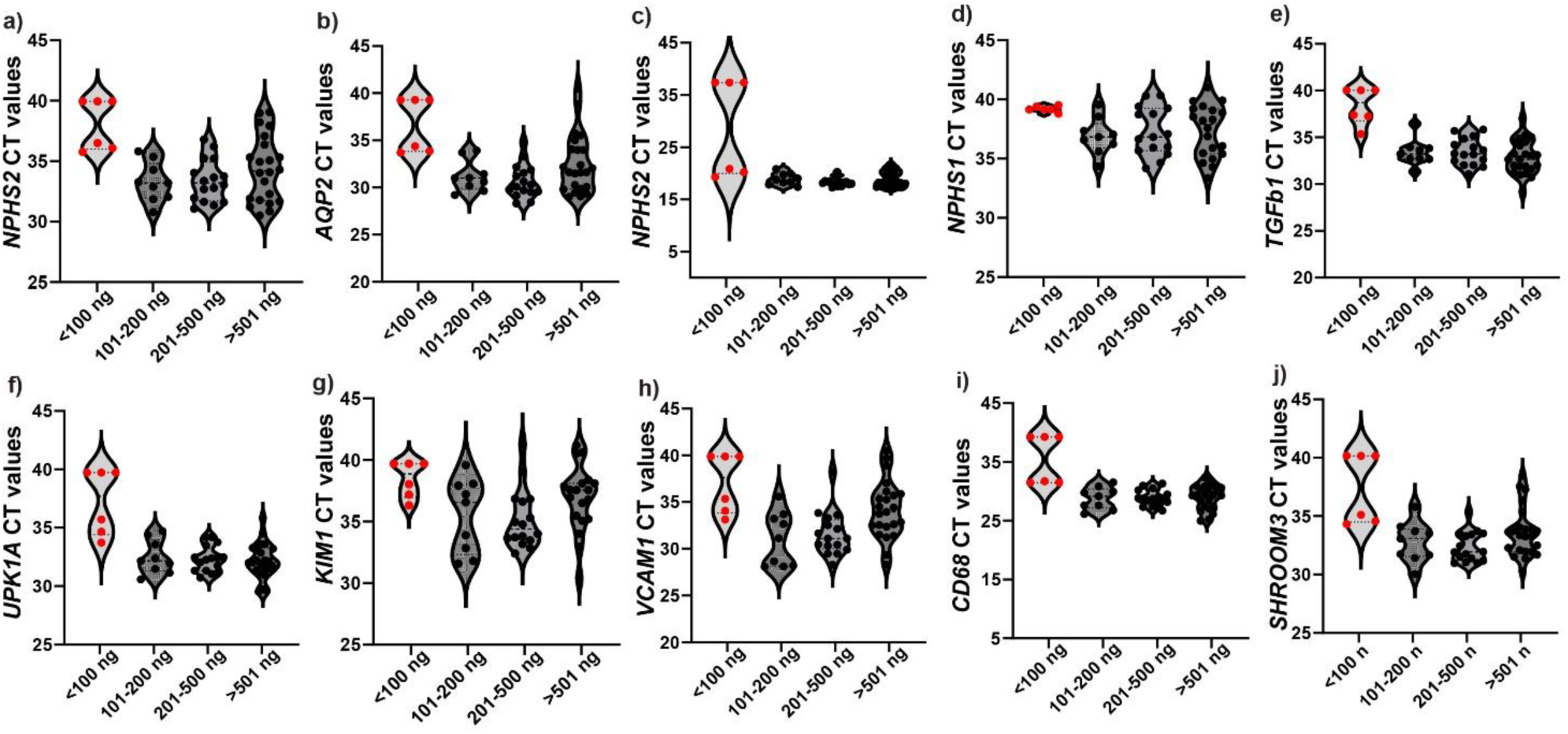
The panel presents CT values for all 10 genes plotted against total RNA concentration from cases. The data indicate that cases with <100 ng RNA had high CT values for most of the genes assessed. The total RNA isolated from the urine pellet ranged from 100 to 1000 ng.

**Supplementary Figure 6:**
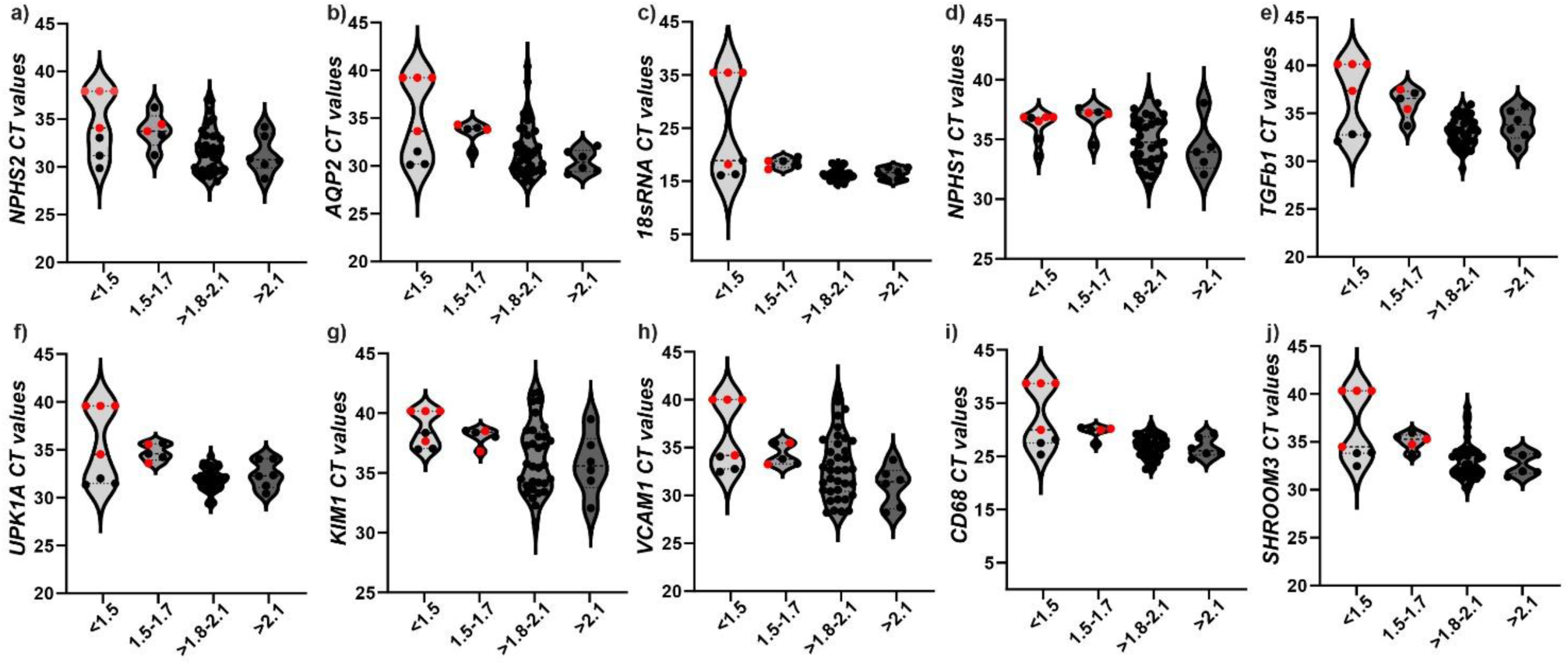
The panel showing CT values for all 10 genes plotted against three RNA purity groups: <1.5, 1.5 to 1.7, >1.8 to 2.1, and >2.1. The data show that low absorbance ratios had an inverse correlation with the CT values of any of the genes assessed, including the housekeeping gene (*18S rRNA*).

**Supplementary Figure 7:**
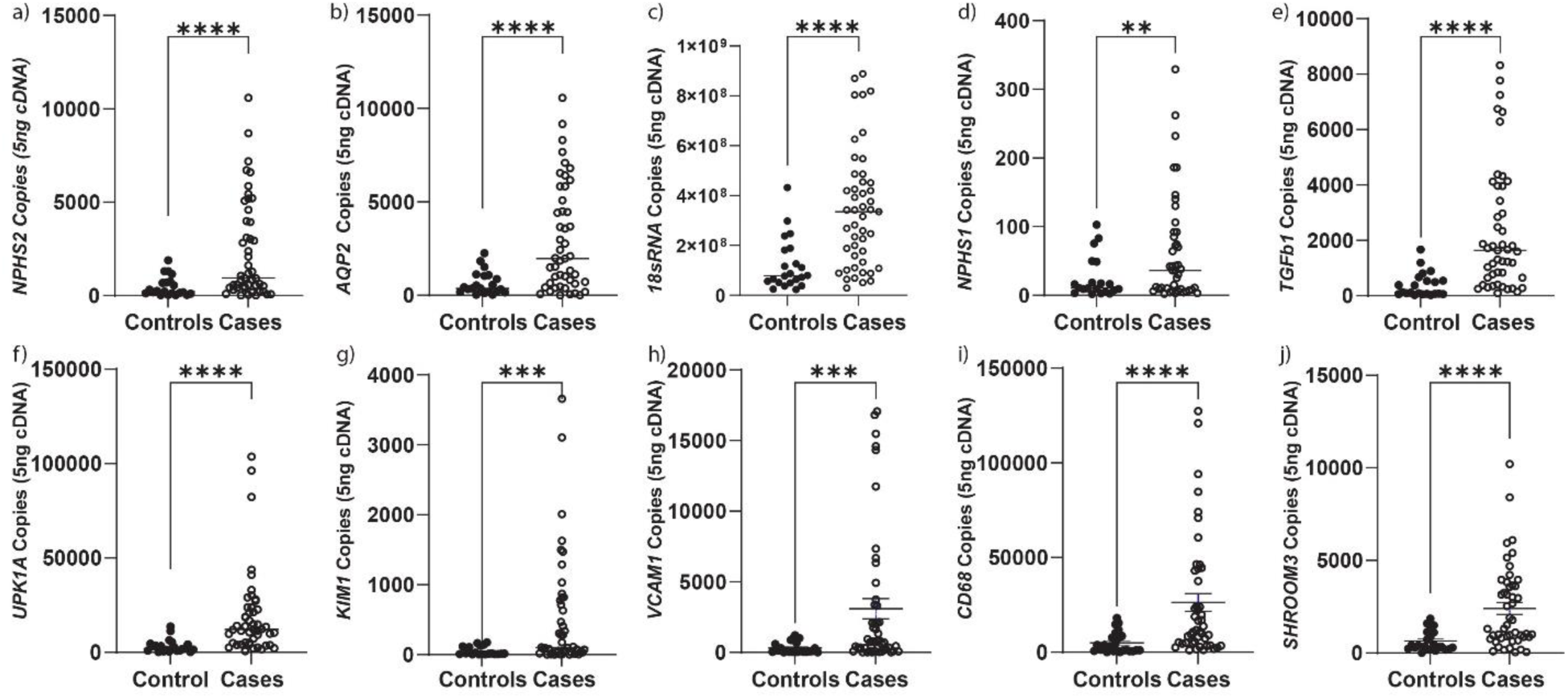
Urinary mRNA biomarker expression comparing raw copy numbers between controls and cases. (a-j) Scatter plots showing mRNA copy numbers (per 5 ng of cDNA) for 10 genes (a) *NPHS2*, (b) *AQP2*, (c) *18s rRNA*, (d) *NPHS1*, (e) *TGFβ1*, (f) *UPK1A*, (g) *KIM1*, (h) *VCAM1*, (i) *CD68*, and (j) *SHROOM3* in control versus pathology groups using unpaired t-test with Welch’s correction. All genes exhibited significantly higher expression in the pathology group compared to controls. Statistical significance is indicated as follows: *p*<0.05, \**p*<0.01, \*\**p*<0.001, \*\*\**p*<0.0001.

**Supplementary Figure 8:**
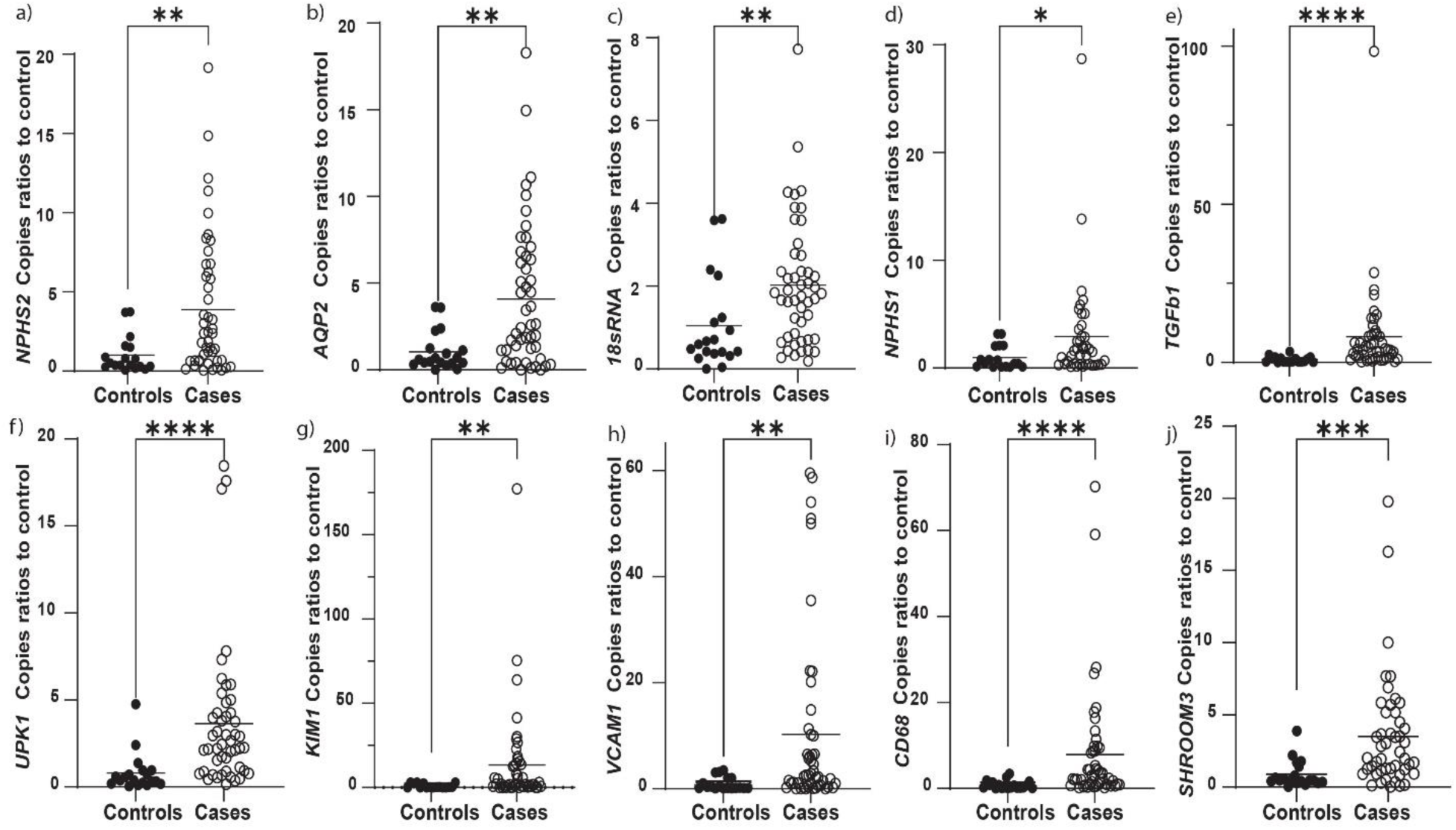
Urinary mRNA biomarker expression comparing fold changes in copy numbers between controls and cases. (a-j) Scatter plots showing mRNA fold changes for 10 genes using mean of controls run in the same plates (a) *NPHS2*, (b) *AQP2*, (c) *18s rRNA*, (d) *NPHS1*, (e) *TGFβ1*, (f) *UPK1A*, (g) *KIM1*, (h) *VCAM1*, (i) *CD68*, and (j) *SHROOM3* in control versus pathology groups using unpaired t-test with Welch’s correction. Similar to raw numbers, all genes exhibited significantly higher expression in the pathology group compared to controls. Statistical significance is indicated as follows: *p*<0.05, \**p*<0.01, \*\**p*<0.001, \*\*\**p*<0.0001.

**Supplementary Figure 9:**
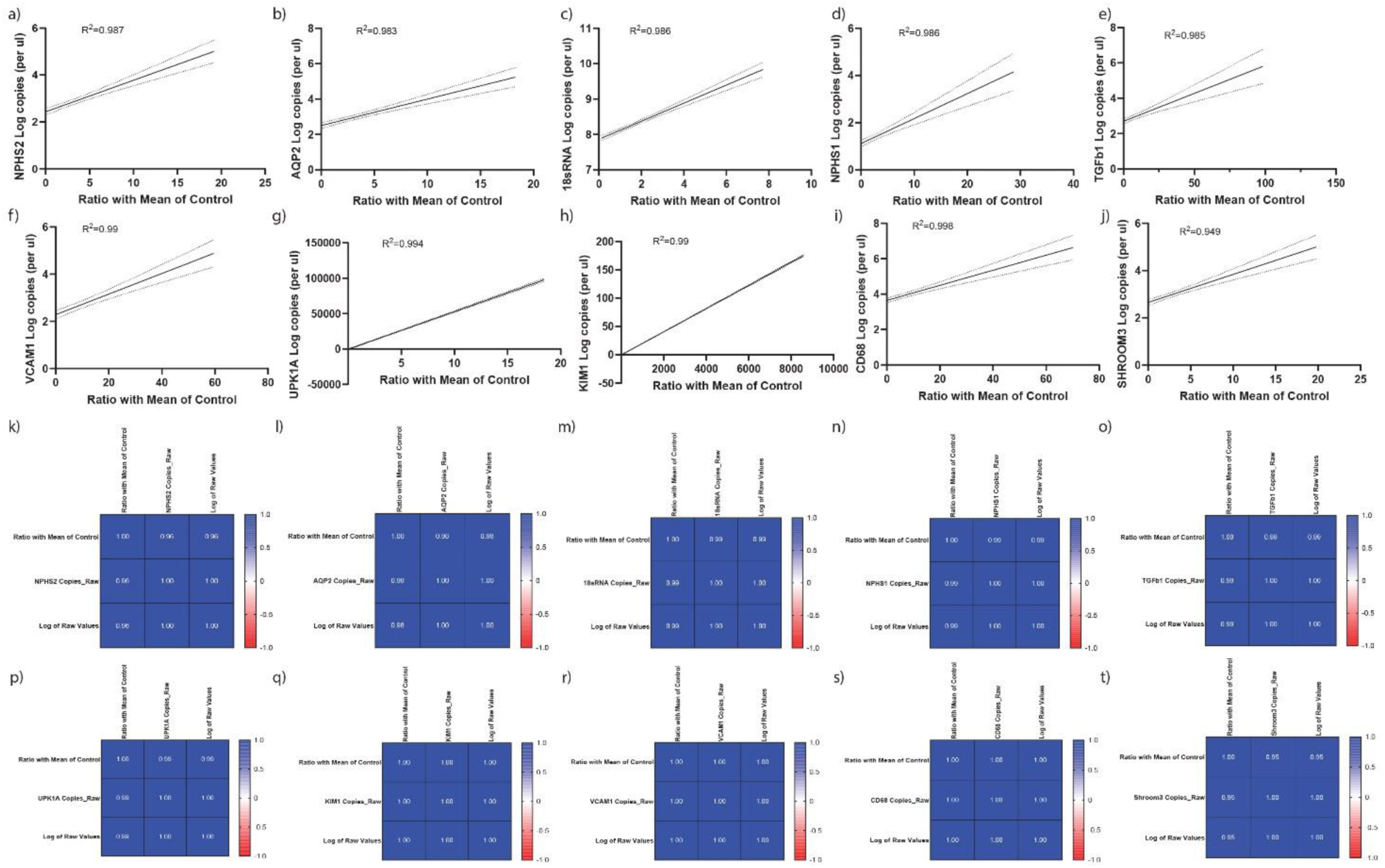
Simple linear regression curves (a-j) showing the relationship between the log-transformed copy numbers and the ratio calculated by averaging the control values for all 10 genes. Correlation matrix (k-t) showing the relationships between the ratio of the mean control to raw copy numbers and the log-transformed raw copy numbers. Strong correlations are observed between these values for all 10 genes.

**Supplementary Figure 10:**
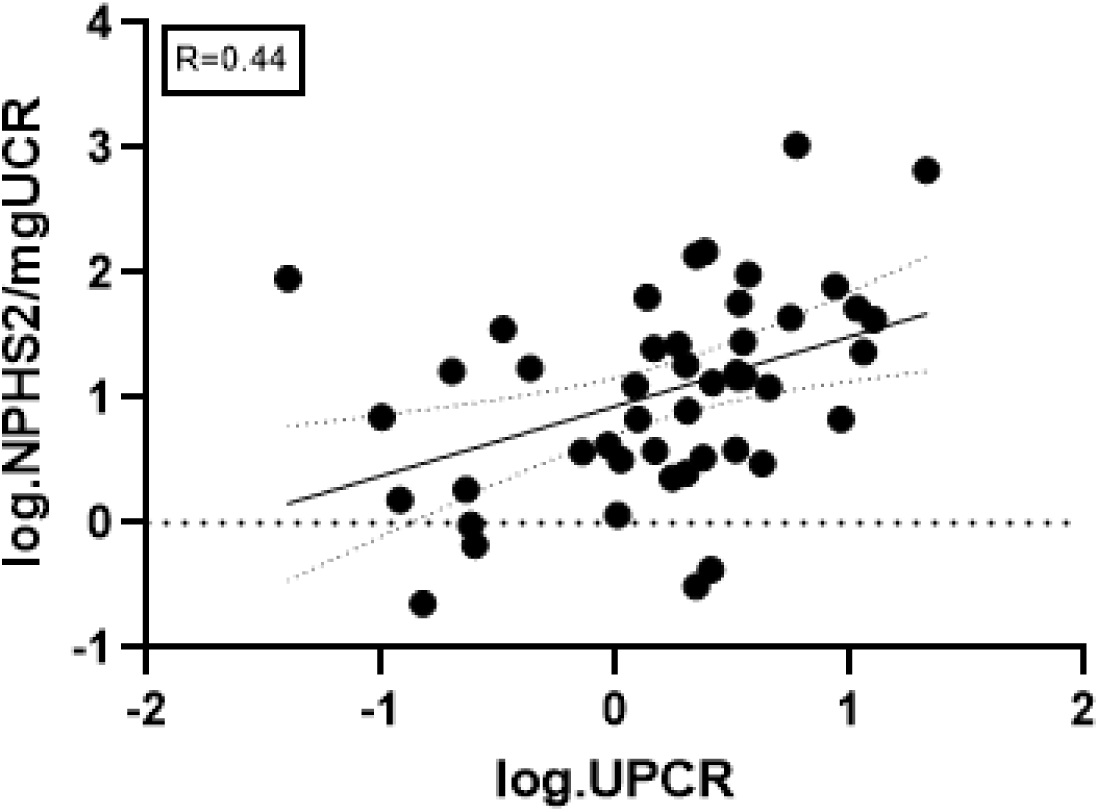
Simple linear regression curve showing the correlation between the log of NPHS2 normalised by UCR and the log of UPCR.

**Supplementary Table-1:**
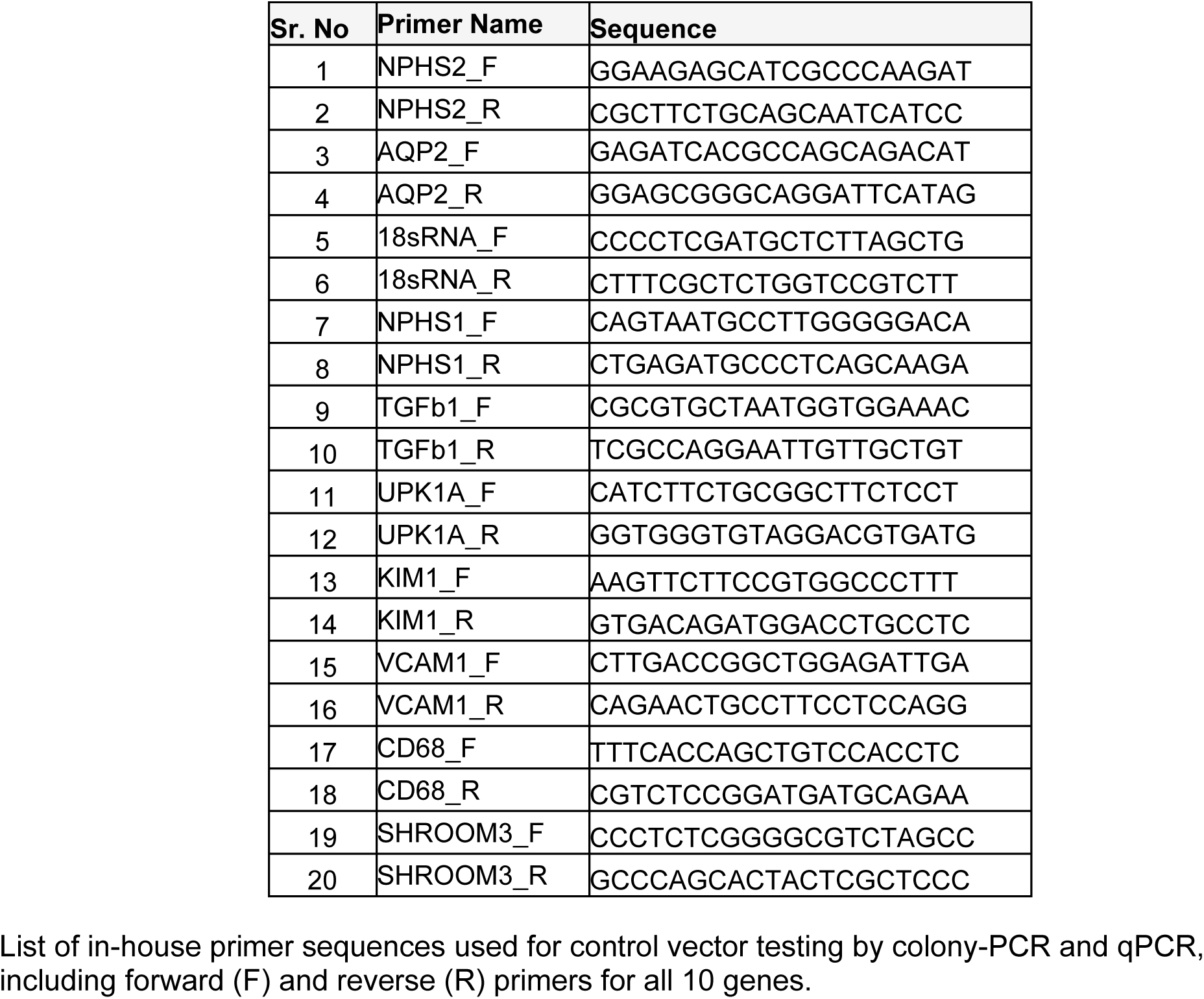
In-house primers used for control vector testing by colony-PCR and qPCR.

**Supplementary Table-2:** Table showing TaqMan assay probes used in 10 genes of NephroDx.

| Sr. Number | Gene Name | TaqMan Probes Assay ID | Panel Name | Company |
| --- | --- | --- | --- | --- |
| 1 | *NPHS2* | Hs00922492_m1 | Panel-1 | Applied Biosystems |
| 2 | *AQP2* | Hs00166640_m1 |  |  |
| 3 | *18s rRNA* | Hs03928985_g1 |  |  |
| 4 | *NPHS1* | Hs00190446_m1 | Panel-2 |  |
| 5 | *TGFB1* | Hs00171257_1ml |  |  |
| 6 | *UPK1A* | Hs00199638_m1 |  |  |
| 7 | *KIM1(HAVCR1)* | Hs00273334_m1 | Panel-3 |  |
| 8 | *VCAM1* | Hs01003372_m1 |  |  |
| 9 | *CD68* | Hs00154355_m1 |  |  |
| 10 | *SHROOM3* | Hs01068190_m1 | Individual Gene Assay |  |

TaqMan assay probes used for the 10 genes, with corresponding assay IDs, panel assignments, and supplier (Applied Biosystems).

**Supplementary Table-3:** Pathological and clinical data.

| Clinical data |  |  |  |  |
| --- | --- | --- | --- | --- |
|  | Gene Name | Correlation Coefficient | P-Value | Adjusted P-Value |
| Dipstick proteinuria (at Biopsy) | *CD68* | 0.44 | <0.01 | 0.01 |
|  | *NPHS1* | 0.43 | <0.01 | 0.02 |
|  | *NPHS2* | 0.40 | <0.01 | 0.025 |
| Pathological data |  |  |  |  |
|  | Digital Pathology Feature | Correlation Coefficient | P-Value | Adjusted P-Value |
| Calculated Tubular Score | % of abnormal interstitial area | -0.63 | <0.01 | 0.005 |
|  | % of atrophic tubules | -0.58 | <0.01 | 0.005 |
|  | Severity of atrophy within tubules | -0.36 | <0.01 | 0.005 |
|  | Number of normal tubules | 0.55 | <0.01 | 0.005 |
|  | Concomitant eGFR | 0.59 | <0.01 | 0.005 |
| IF/TA Score (Mild, Moderate, Severe) | % of abnormal interstitial area | 0.51 | <0.01 | 0.005 |
|  | % of atrophic tubules | 0.47 | <0.01 | 0.005 |
|  | Number of atrophic tubules | 0.46 | <0.01 | 0.005 |

Spearman correlations of clinical data (dipstick proteinuria) with gene expression, and pathological features (digital pathology metrics, IF/TA score, eGFR), showing correlation coefficients.

**Supplementary Table-4:**
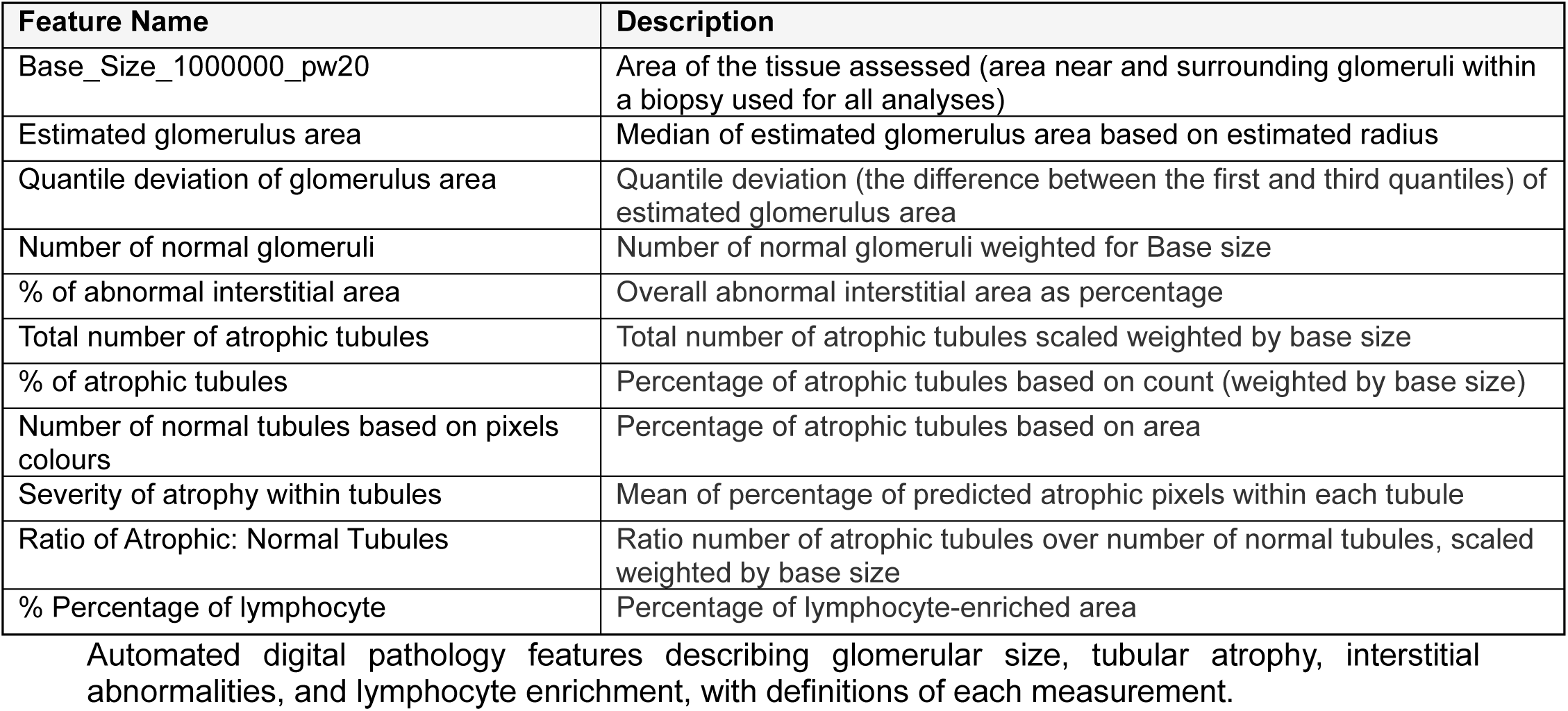
Automated Digital Pathology Features and Descriptions.

## References

1. Go AS, Chertow GM, Fan D, McCulloch CE, Hsu CY. Chronic kidney disease and the risks of death, cardiovascular events, and hospitalization. N Engl J Med. 2004;351(13):1296–305.

2. Chronic Kidney Disease Prognosis C, Matsushita K, van der Velde M, Astor BC, Woodward M, Levey AS, et al. Association of estimated glomerular filtration rate and albuminuria with all-cause and cardiovascular mortality in general population cohorts: a collaborative meta-analysis. Lancet. 2010;375(9731):2073–81.

3. Grams ME, Sang Y, Ballew SH, Carrero JJ, Djurdjev O, Heerspink HJL, et al. Predicting timing of clinical outcomes in patients with chronic kidney disease and severely decreased glomerular filtration rate. Kidney Int. 2018;93(6):1442–51.

4. Lopez-Giacoman S, Madero M. Biomarkers in chronic kidney disease, from kidney function to kidney damage. World J Nephrol. 2015;4(1):57–73.

5. Kidney Disease: Improving Global Outcomes CKDWG. KDIGO 2024 Clinical Practice Guideline for the Evaluation and Management of Chronic Kidney Disease. Kidney Int. 2024;105(4S):S117–S314.

6. Hickson LJ, Chaudhary S, Williams AW, Dillon JJ, Norby SM, Gregoire JR, et al. Predictors of outpatient kidney function recovery among patients who initiate hemodialysis in the hospital. Am J Kidney Dis. 2015;65(4):592–602.

7. Malhotra R, Craven T, Ambrosius WT, Killeen AA, Haley WE, Cheung AK, et al. Effects of Intensive Blood Pressure Lowering on Kidney Tubule Injury in CKD: A Longitudinal Subgroup Analysis in SPRINT. Am J Kidney Dis. 2019;73(1):21–30.

8. Peterson JC, Adler S, Burkart JM, Greene T, Hebert LA, Hunsicker LG, et al. Blood pressure control, proteinuria, and the progression of renal disease. The Modification of Diet in Renal Disease Study. Ann Intern Med. 1995;123(10):754–62.

9. Hogg RJ, Portman RJ, Milliner D, Lemley KV, Eddy A, Ingelfinger J. Evaluation and management of proteinuria and nephrotic syndrome in children: recommendations from a pediatric nephrology panel established at the National Kidney Foundation conference on proteinuria, albuminuria, risk, assessment, detection, and elimination (PARADE). Pediatrics. 2000;105(6):1242–9.

10. Waikar SS, Betensky RA, Emerson SC, Bonventre JV. Imperfect gold standards for kidney injury biomarker evaluation. J Am Soc Nephrol. 2012;23(1):13–21.

11. Waikar SS, Bonventre JV. Creatinine kinetics and the definition of acute kidney injury. J Am Soc Nephrol. 2009;20(3):672–9.

12. Hogan JJ, Mocanu M, Berns JS. The Native Kidney Biopsy: Update and Evidence for Best Practice. Clin J Am Soc Nephrol. 2016;11(2):354–62.

13. Lake BB, Menon R, Winfree S, Hu Q, Melo Ferreira R, Kalhor K, et al. An atlas of healthy and injured cell states and niches in the human kidney. Nature. 2023;619(7970):585–94.

14. Park J, Liu CL, Kim J, Susztak K. Understanding the kidney one cell at a time. Kidney Int. 2019;96(4):862–70.

15. Luciano RL, Moeckel GW. Update on the Native Kidney Biopsy: Core Curriculum 2019. Am J Kidney Dis. 2019;73(3):404–15.

16. Brachemi S, Bollee G. Renal biopsy practice: What is the gold standard? World J Nephrol. 2014;3(4):287–94.

17. Sato Y, Wharram BL, Lee SK, Wickman L, Goyal M, Venkatareddy M, et al. Urine podocyte mRNAs mark progression of renal disease. J Am Soc Nephrol. 2009;20(5):1041–52.

18. Suthanthiran M, Schwartz JE, Ding R, Abecassis M, Dadhania D, Samstein B, et al. Urinary-cell mRNA profile and acute cellular rejection in kidney allografts. N Engl J Med. 2013;369(1):20–31.

19. Klocke J, Kim SJ, Skopnik CM, Hinze C, Boltengagen A, Metzke D, et al. Urinary single-cell sequencing captures kidney injury and repair processes in human acute kidney injury. Kidney Int. 2022;102(6):1359–70.

20. Fukuda A, Sato Y, Shibata H, Fujimoto S, Wiggins RC. Urinary podocyte markers of disease activity, therapeutic efficacy, and long-term outcomes in acute and chronic kidney diseases. Clin Exp Nephrol. 2024;28(6):496–504.

21. Wickman L, Afshinnia F, Wang SQ, Yang Y, Wang F, Chowdhury M, et al. Urine podocyte mRNAs, proteinuria, and progression in human glomerular diseases. J Am Soc Nephrol. 2013;24(12):2081–95.

22. Verma A, Muthukumar T, Yang H, Lubetzky M, Cassidy MF, Lee JR, et al. Urinary cell transcriptomics and acute rejection in human kidney allografts. JCI Insight. 2020;5(4).

23. Hinze C, Schmidt-Ott KM. Acute kidney injury biomarkers in the single-cell transcriptomic era. Am J Physiol Cell Physiol. 2022;323(5):C1430–C43.

24. Barsotti GC, Luciano R, Kumar A, Meliambro K, Kakade V, Tokita J, et al. Rationale and Design of a Phase 2, Double-blind, Placebo-Controlled, Randomized Trial Evaluating AMP Kinase-Activation by Metformin in Focal Segmental Glomerulosclerosis. Kidney Int Rep. 2024;9(5):1354–68.

25. Franceschini N, North KE, Kopp JB, McKenzie L, Winkler C. NPHS2 gene, nephrotic syndrome and focal segmental glomerulosclerosis: a HuGE review. Genet Med. 2006;8(2):63–75.

26. Roselli S, Gribouval O, Boute N, Sich M, Benessy F, Attie T, et al. Podocin localizes in the kidney to the slit diaphragm area. Am J Pathol. 2002;160(1):131–9.

27. Schwarz K, Simons M, Reiser J, Saleem MA, Faul C, Kriz W, et al. Podocin, a raft-associated component of the glomerular slit diaphragm, interacts with CD2AP and nephrin. J Clin Invest. 2001;108(11):1621–9.

28. Latt KZ, Heymann J, Jessee JH, Rosenberg AZ, Berthier CC, Arazi A, et al. Urine Single-Cell RNA Sequencing in Focal Segmental Glomerulosclerosis Reveals Inflammatory Signatures. Kidney Int Rep. 2022;7(2):289–304.

29. Boute N, Gribouval O, Roselli S, Benessy F, Lee H, Fuchshuber A, et al. NPHS2, encoding the glomerular protein podocin, is mutated in autosomal recessive steroid-resistant nephrotic syndrome. Nat Genet. 2000;24(4):349–54.

30. Kestila M, Lenkkeri U, Mannikko M, Lamerdin J, McCready P, Putaala H, et al. Positionally cloned gene for a novel glomerular protein--nephrin--is mutated in congenital nephrotic syndrome. Mol Cell. 1998;1(4):575–82.

31. Khoshnoodi J, Sigmundsson K, Ofverstedt LG, Skoglund U, Obrink B, Wartiovaara J, et al. Nephrin promotes cell-cell adhesion through homophilic interactions. Am J Pathol. 2003;163(6):2337–46.

32. Fukuda A, Minakawa A, Kikuchi M, Sato Y, Nagatomo M, Nakamura S, et al. Urinary podocyte mRNAs precede microalbuminuria as a progression risk marker in human type 2 diabetic nephropathy. Sci Rep. 2020;10(1):18209.

33. Karmakova capital Te CAC, Sergeeva NS, Kanukoev capital Ka C, Alekseev BY, Kaprin capital A C. Kidney Injury Molecule 1 (KIM-1): a Multifunctional Glycoprotein and Biological Marker (Review). Sovrem Tekhnologii Med. 2021;13(3):64–78.

34. Han WK, Bailly V, Abichandani R, Thadhani R, Bonventre JV. Kidney Injury Molecule-1 (KIM-1): a novel biomarker for human renal proximal tubule injury. Kidney Int. 2002;62(1):237–44.

35. Fushimi K, Uchida S, Hara Y, Hirata Y, Marumo F, Sasaki S. Cloning and expression of apical membrane water channel of rat kidney collecting tubule. Nature. 1993;361(6412):549–52.

36. Nielsen S, DiGiovanni SR, Christensen EI, Knepper MA, Harris HW. Cellular and subcellular immunolocalization of vasopressin-regulated water channel in rat kidney. Proc Natl Acad Sci U S A. 1993;90(24):11663–7.

37. Liao J, Yu Z, Chen Y, Bao M, Zou C, Zhang H, et al. Single-cell RNA sequencing of human kidney. Sci Data. 2020;7(1):4.

38. Chung S, Overstreet JM, Li Y, Wang Y, Niu A, Wang S, et al. TGF-beta promotes fibrosis after severe acute kidney injury by enhancing renal macrophage infiltration. JCI Insight. 2018;3(21).

39. Kanai H, Mitsuhashi H, Ono K, Yano S, Naruse T. Increased excretion of urinary transforming growth factor beta in patients with focal glomerular sclerosis. Nephron. 1994;66(4):391–5.

40. Fukuda A, Wickman LT, Venkatareddy MP, Sato Y, Chowdhury MA, Wang SQ, et al. Angiotensin II-dependent persistent podocyte loss from destabilized glomeruli causes progression of end stage kidney disease. Kidney Int. 2012;81(1):40–55.

41. Segerer S, Heller F, Lindenmeyer MT, Schmid H, Cohen CD, Draganovici D, et al. Compartment specific expression of dendritic cell markers in human glomerulonephritis. Kidney Int. 2008;74(1):37–46.

42. Yamamoto K, Oda T, Uchida T, Takechi H, Oshima N, Kumagai H. Evaluating the State of Glomerular Disease by Analyzing Urinary Sediments: mRNA Levels and Immunofluorescence Staining for Various Markers. Int J Mol Sci. 2024;25(2).

43. Xu L, Guo J, Moledina DG, Cantley LG. Immune-mediated tubule atrophy promotes acute kidney injury to chronic kidney disease transition. Nat Commun. 2022;13(1):4892.

44. Ellis JW, Chen MH, Foster MC, Liu CT, Larson MG, de Boer I, et al. Validated SNPs for eGFR and their associations with albuminuria. Hum Mol Genet. 2012;21(14):3293–8.

45. Paul A, Lawlor A, Cunanan K, Gaheer PS, Kalra A, Napoleone M, et al. The Good and the Bad of SHROOM3 in Kidney Development and Disease: A Narrative Review. Can J Kidney Health Dis. 2023;10:20543581231212038.

46. Prokop JW, Yeo NC, Ottmann C, Chhetri SB, Florus KL, Ross EJ, et al. Characterization of Coding/Noncoding Variants for SHROOM3 in Patients with CKD. J Am Soc Nephrol. 2018.

47. Menon MC, Chuang PY, Li Z, Wei C, Zhang W, Luan Y, et al. Intronic locus determines SHROOM3 expression and potentiates renal allograft fibrosis. J Clin Invest. 2014.

48. Wei C, Banu K, Garzon F, Basgen JM, Philippe N, Yi Z, et al. SHROOM3-FYN Interaction Regulates Nephrin Phosphorylation and Affects Albuminuria in Allografts. J Am Soc Nephrol. 2018;29(11):2641–57.

49. Reghuvaran A, Kumar A, Lin Q, Rajeevan N, Sun Z, Shi H, et al. Shroom3-Rock interaction and profibrotic function: Resolving mechanism of an intronic CKD risk allele. bioRxiv. 2024.

50. Carpenter AR, Becknell MB, Ching CB, Cuaresma EJ, Chen X, Hains DS, et al. Uroplakin 1b is critical in urinary tract development and urothelial differentiation and homeostasis. Kidney Int. 2016;89(3):612–24.

51. Fink EE, Sona S, Tran U, Desprez PE, Bradley M, Qiu H, et al. Single-cell and spatial mapping Identify cell types and signaling Networks in the human ureter. Dev Cell. 2022;57(15):1899–916 e6.

52. Galichon P, Amrouche L, Hertig A, Brocheriou I, Rabant M, Xu-Dubois YC, et al. Urinary mRNA for the Diagnosis of Renal Allograft Rejection: The Issue of Normalization. Am J Transplant. 2016;16(10):3033–40.

53. Yi Z, Salem F, Menon MC, Keung K, Xi C, Hultin S, et al. Deep learning identified pathological abnormalities predictive of graft loss in kidney transplant biopsies. Kidney Int. 2022;101(2):288–98.

54. Klötzer KA, Abedini A, Li S, Balzer MS, Liang X, Levinsohn J, et al. Analysis of individual patient pathway coordination in a cross-species single-cell kidney atlas. Nature Genetics. 2025:1–13.

55. Abedini A, Levinsohn J, Klötzer KA, Dumoulin B, Ma Z, Frederick J, et al. Single-cell multi-omic and spatial profiling of human kidneys implicates the fibrotic microenvironment in kidney disease progression. Nature genetics. 2024;56(8):1712–24.

56. Abedini A, Zhu YO, Chatterjee S, Halasz G, Devalaraja-Narashimha K, Shrestha R, et al. Urinary Single-Cell Profiling Captures the Cellular Diversity of the Kidney. J Am Soc Nephrol. 2021;32(3):614–27.

57. Fogazzi GB. Crystalluria: a neglected aspect of urinary sediment analysis. Nephrol Dial Transplant. 1996;11(2):379–87.

58. Saetun P, Semangoen T, Thongboonkerd V. Characterizations of urinary sediments precipitated after freezing and their effects on urinary protein and chemical analyses. Am J Physiol Renal Physiol. 2009;296(6):F1346–54.

59. Ackerman AL, Anger JT, Khalique MU, Ackerman JE, Tang J, Kim J, et al. Optimization of DNA extraction from human urinary samples for mycobiome community profiling. PLoS One. 2019;14(4):e0210306.

60. Medeiros M, Sharma VK, Ding R, Yamaji K, Li B, Muthukumar T, et al. Optimization of RNA yield, purity and mRNA copy number by treatment of urine cell pellets with RNAlater. J Immunol Methods. 2003;279(1-2):135–42.

61. Landini G, Martinelli G, Piccinini F. Colour deconvolution: stain unmixing in histological imaging. Bioinformatics. 2021;37(10):1485–7.

62. John M Basgen ACR, Qisheng Lin, Khadija Banu, Hongmei Shi, John Pell, Jenna DiRito, Gregory T Tietjen, Sudhir Perincheri, Dennis G Moledina, Francis Perry Wilson, Madhav C Menon*. Comparative evaluation of glomerular morphometric techniques reveals differential technical artefacts between FSGS and normal glomeruli BioRxiv: Cold Spring Harbor Laboratory; 2022 [Available from: https://www.biorxiv.org/content/10.1101/2022.02.15.480420v1.

63. Ichimura T, Brooks CR, Bonventre JV. Kim-1/Tim-1 and immune cells: shifting sands. Kidney Int. 2012;81(9):809–11.

64. Kottgen A, Pattaro C, Boger CA, Fuchsberger C, Olden M, Glazer NL, et al. New loci associated with kidney function and chronic kidney disease. Nat Genet. 2010;42(5):376–84.

65. Loeb GB, Kathail P, Shuai RW, Chung R, Grona RJ, Peddada S, et al. Variants in tubule epithelial regulatory elements mediate most heritable differences in human kidney function. Nat Genet. 2024.

